# Deep learning of fundus and optical coherence tomography images enables identification of diverse genetic and environmental factors associated with eye aging

**DOI:** 10.1101/2021.06.24.21259471

**Authors:** Alan Le Goallec, Samuel Diai, Sasha Collin, Théo Vincent, Chirag J. Patel

**Author notes:** Corresponding author, **Contact information:** Chirag J Patel. Co-second authors (Samuel Diai, Sasha Collin).

## Abstract

**Background:** The rate at which different portions of the eye ages can be measured using eye fungus and optical coherence tomography (OCT) images; however, their genetic and environmental contributors have been elusive.

**Methods:** We built an eye age predictor by training convolutional neural networks to predict age from 175,000 eye fundus and OCT images from participants of the UK Biobank cohort, capturing two different dimensions of eye (retinal, macula, fovea) aging. We performed a genome-wide association study (GWAS) and high-throughput epidemiology to identify novel genetic and environmental variables associated with the new age predictor, finding variables associated with accelerated eye aging.

**Findings:** Fundus-based and OCT-based eye aging capture different dimensions of eye aging, whose combination predicted chronological age with an R^2^ and mean absolute error of 83.6±0.6%/2.62±0.05 years. In comparison, the fundus-based and OCT-based predictor alone predicted age with R^2^ of 76.6±1.3% vs. 70.8±1.2% respectively. Accelerated eye fundus- and OCT-measured accelerated aging has a significant genetic component, with heritability (total contribution of GWAS variants) of 26 and 23% respectively. For eye fundus measured aging, we report novel variants in the *FAM150B* gene (*ALKAL2*, or *ALK* ligand 2) (p<1×10^-150^); for OCT-measured eye aging, we found variants in genes such as *CFH* (complement factor H), *COL4A4* (type 4 collagen), and *RLBP* (retinaldehyde binding protein 1, all p<1×10^-20^). Eye accelerated aging is also associated with behaviors and socioeconomic status, such as sleep deprivation and lower income.

**Conclusions:** Our new deep-learning-based digital readouts, the best eye aging predictor to date, suggest a biological basis of eye aging. These new data can be harnessed for scalable genetic and epidemiological dissection and discovery of aging specific to different components of the eye and their relationship with different diseases of aging.

**Funding:** National Institutes of Health, National Science Foundation, MassCATS, Sanofi. Funders had no role in the project.

**Research in context:** 

**Evidence before this study:** We performed a search on NCBI PubMed and Google Scholar searching for the terms, “eye aging”, “optical coherence tomography” (OCT), “fundus”, and/or “deep learning”. We found others have shown feasibility of predicting chronological age from eye image modalities, finding five publications that demonstrated chronological age may be predicted from images inside and outside of the eye, with mean absolute errors ranging from 2.3-5.82 years.

**Added value of this study:** Our new eye age predictor combines both OCT and fundus images to assemble the most accurate fundus/OCT age predictor to date (mean absolute error of 2.62 years). Second, we have identified new genetic loci (e.g., in *FAM150B*) and epidemiological associations with eye accelerated age, highlighting the biological and environmental correlates of eye age, elusive in other investigations and made scalable by deep learning.

## Introduction

Aging affects all components of the eye (eyelids, lacrimal system, cornea, trabecular mesh work, uvea, crystalline lens, retina, macula (Salvi et al., 2006)) and is associated with the onset of age-related eye diseases such as presbyopia, cataract, glaucoma, macular degeneration and retinopathy (Visser, 2006) leading to reduced quality of life, increased healthcare costs (Rein et al., 2006), and shortened lifespan due to the increased risk for falls (Lord et al., 2010).

Age predictors have recently been derived to measure accelerated aging(Horvath, 2013; Le Goallec et al., 2022) and of the eye (Poplin et al., 2018). Here, inspired by these approaches, we use machine learning algorithms to predict the age of the participant (also referred to as “chronological age”) from fundus and optical coherence tomography (OCT) images of the eye. Fundus images capture the rear of the eye, or fundus, along with the retina and macula. OCT images a cross-sectional image of the retina, a different dimension of fundus images. The predictions of the model are interpreted as the participant’s eye age (also referred to as “biological age”). We next can define accelerated eye aging as the difference between eye age and chronological age. Age has for example already been predicted from fundus(Poplin et al., 2018), optical coherence tomography (Chueh et al., n.d.; Shigueoka et al., 2021), iris (Erbilek et al., 2013; Rajput and Sable, n.d.; Sgroi et al., 2013) and eye corner images (Bobrov et al., 2018). Identifying the genetic and non-genetic factors underlying different physiological measure of eye aging has not, hitherto, been done.

In the following, we leverage 175,000 OCT and eye fundus images (Figure 1B), along with eye biomarkers (acuity, refractive error, intraocular pressure) collected from 37-82 year-old UK Biobank (Sudlow et al., 2015) participants (Figure 1A), and use deep learning to build eye age predictors. We perform a genome wide association study [GWAS] and an X-wide association study [XWAS] to identify genetic and non-genetic factors (e.g biomarkers, phenotypes, diseases, environmental and socioeconomic variables) associated with eye aging. (Figure 1C)

**Figure 1:**
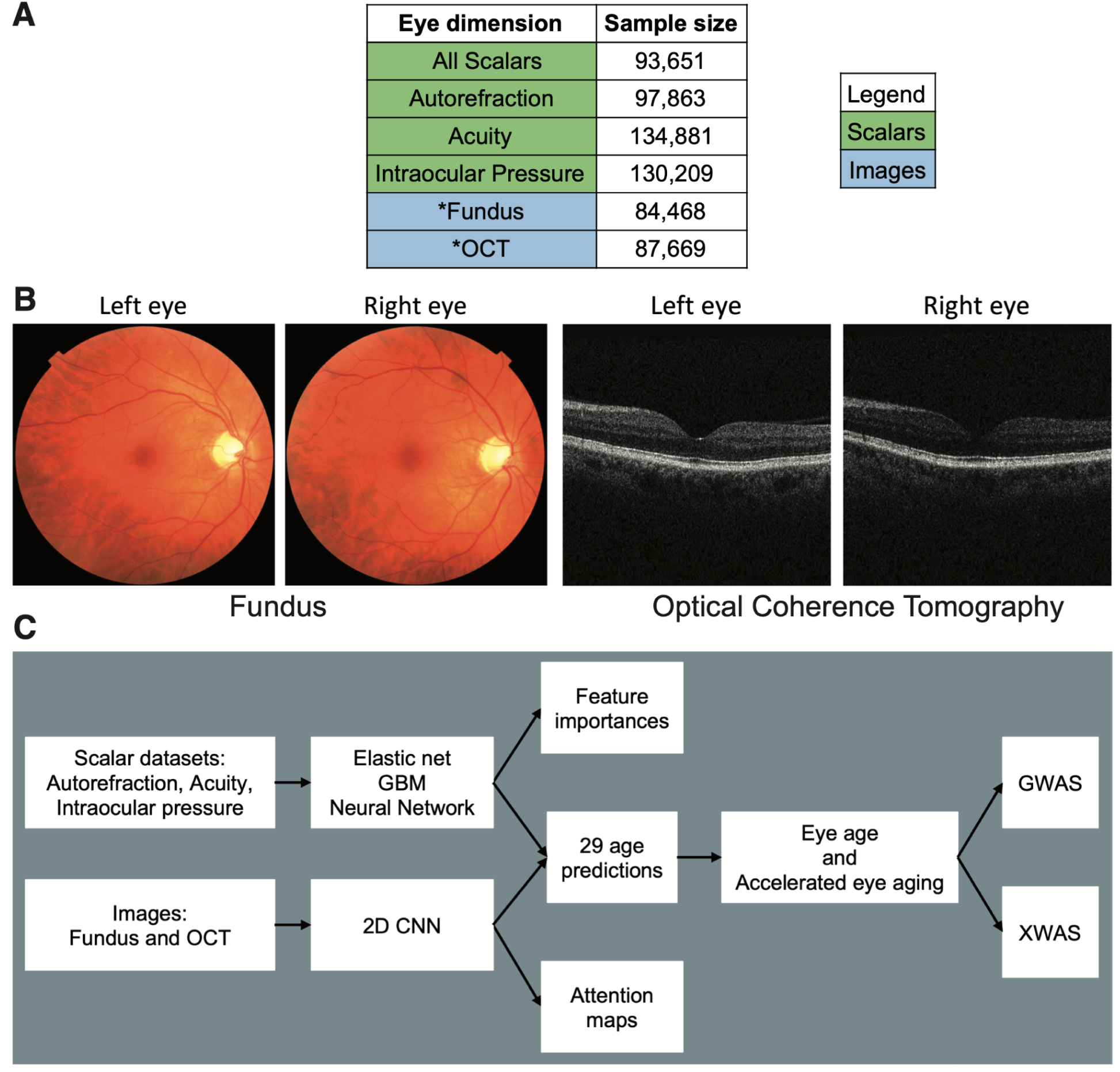
Overview of the datasets and analytic pipeline. A - Eye aging dimensions. B - Sample eye fundus and optical coherence tomography images. C - Analytic pipeline. Legend for A - * Correspond to eye dimensions for which we performed a GWAS and XWAS analysis

## Methods

### Data and materials availability

The anonymous data are available through the UK Biobank (project ID: 52887). The code can be found at https://github.com/Deep-Learning-and-Aging. We will make the biological age phenotypes available through UK Biobank upon publication. The results can be interactively and extensively explored at https://www.multidimensionality-of-aging.net/, a website where we display and compare the performance and properties of the different biological age predictors we built. Select “Eyes” as the aging dimension to view. The GWAS results can be found at via FUMA: OCT: https://fuma.ctglab.nl/browse/495; Fundus: https://fuma.ctglab.nl/browse/494. Our code can be found at https://github.com/Deep-Learning-and-Aging. The supplementary tables can be found here: https://www.dropbox.com/s/px2re5qw9n11htt/Supplementary%20data.zip?dl=0

### Cohort Dataset: Participants of the UK Biobank

We leveraged the UK Biobank(Sudlow et al., 2015) cohort (project ID: 52887). The UKB cohort consists of data originating from a large biobank collected from 502,211 de-identified participants in the United Kingdom that were aged between 37 years and 74 years at enrollment (starting in 2006). Out of these participants, approximately 87,000 had fundus and OCT images collected from them. The Harvard internal review board (IRB) deemed the research as non-human subjects research (IRB: IRB16-2145).

### Data types and Preprocessing

The data preprocessing step is different for the different data modalities: demographic variables, scalar predictors and images. We define scalar predictors as predictors whose information can be encoded in a single number, such as eye spherical power, as opposed to data with a higher number of dimensions such as images (two dimensions, which are the height and the width of the image). Please see Supplementary Methods/Figures for detailed pre-processing of the demographic, scalar, and eye fundus/OCT images (Figure S5, Figure S6, Figure S7, Figure S8, Figure S9, Figure S10, Figure S11). For scalar datasets, we used elastic nets, gradient boosted machines [GBMs] and fully connected neural networks. For images we used two-dimensional convolutional neural networks. For details on algorithms, parameters, and study design of the training tuning and prediction, please refer to the Supplementary Methods.

### Training, tuning and predictions and the interpretability of models

We split the entire dataset into ten data folds. We then tuned the models built on scalar data and the models built on images using two different pipelines. For scalar data-based models, we performed a nested-cross validation. For images-based models, we manually tuned some of the hyperparameters before performing a simple cross-validation. We describe the splitting of the data into different folds and the tuning procedures in detail in the Supplementary Methods.

### Ensembling to improve prediction and define aging dimensions

We built a two-level hierarchy of ensemble models to improve prediction accuracies. At the lowest level, we combined the predictions from different algorithms on the same aging subdimension. For example, we combined the predictions generated by the elastic net, the gradient boosted machine and the neural network from the eye acuity scalar biomarkers. At the second level, we combined the predictions from the different eye dimensions into a general eye age prediction. The ensemble models from the lower levels are hierarchically used as components of the ensemble models of the higher models. For example, the ensemble model built by combining the algorithms trained on eye acuity variables is leveraged when building the general eye aging ensemble model.

We built each ensemble model separately on each of the ten data folds. For example, to build the ensemble model on the testing predictions of the data fold #1, we trained and tuned an elastic net on the validation predictions from the data fold #0 using a 10-folds inner cross-validation, as the validation predictions on fold #0 and the testing predictions on fold #1 are generated by the same model (see Supplementary Methods - Training, tuning and predictions - Images - Scalar data - Nested cross-validation; Supplementary Methods - Training, tuning and predictions - Images - Cross-validation). We used the same hyperparameters space and Bayesian hyperparameters optimization method as we did for the inner cross-validation we performed during the tuning of the non-ensemble models.

To summarize, the testing ensemble predictions are computed by concatenating the testing predictions generated by ten different elastic nets, each of which was trained and tuned using a 10-folds inner cross-validation on one validation data fold (10% of the full dataset) and tested on one testing fold. This is different from the inner-cross validation performed when training the non-ensemble models, which was performed on the “training+validation” data folds, so on 9 data folds (90% of the dataset).

### Evaluating the performance of models

We evaluated the performance of the models using two different metrics: R-Squared [R^2^] and root mean squared error [RMSE]. We computed a confidence interval on the performance metrics in two different ways. First, we computed the standard deviation between the different data folds. The test predictions on each of the ten data folds are generated by ten different models, so this measure of standard deviation captures both model variability and the variability in prediction accuracy between samples. Second, we computed the standard deviation by bootstrapping the computation of the performance metrics 1,000 times. This second measure of variation does not capture model variability but evaluates the variance in the prediction accuracy between samples.

### Eye age definition

We defined the eye age of participants for a specific eye dimension as the prediction outputted by the model trained on the corresponding dataset, after correcting for the bias in the residuals. For each model, participants on the older end of the chronological age distribution tend to be predicted younger than they are. Symmetrically, participants on the younger end of the chronological age distribution tend to be predicted older than they are. This bias does not seem to be biologically driven (Le Goallec et al., 2022). Rather it seems to be statistically driven, as the same 60-year-old individual will tend to be predicted younger in a cohort with an age range of 60-80 years, and to be predicted older in a cohort with an age range of 60-80. We ran a linear regression on the residuals as a function of age for each model and used it to correct each prediction for this statistical bias.

After defining biological age as the corrected prediction, we defined accelerated aging as the corrected residuals. For example, a 60-year-old whose eye fundus images predicted an age of 70 years old after is estimated to have an eye age of 70 years, and an accelerated eye aging of ten years. We emphasize that the step of correction of the predictions and the residuals takes place after the evaluation of the performance of the models but precedes the analysis of the eye ages properties, such as the GWASs.

### Genome-wide association of accelerated eye aging

The UKB contains genome-wide genetic data for 488,251 of the 502,492 participants(Bycroft et al., 2017) under the hg19/GRCh37 build and performed genome-wide association study analysis using using BOLT-LMM (Loh et al., 2018, 2015b) and BOLT-REML (Loh et al., 2015a). For more details, please see the Supplementary Methods.

### Non-genetic correlates of accelerated eye aging

We identified non-genetically measured (i.e factors not measured on a GWAS array) correlates of each aging dimension, which we classified in six categories: biomarkers, clinical phenotypes, diseases, family history, environmental, and socioeconomic variables. We refer to the union of these association analyses as an X-Wide Association Study [XWAS] (Patel et al., 2010). See Supplementary Methods.

## Results

### Chronological age prediction within three years

We leveraged the UK Biobank, a dataset containing 175,000 eye fundus and OCT images (both left and right eyes, so approximately half fewer samples, Figure 1A and B), as well as eye biomarkers (acuity, intraocular and autorefraction tests) collected from 97,000-135,000 (Figure 1A and B) participants aged 37-82 years (Figure S1). We predicted chronological age from images using convolutional neural networks and from scalar biomarkers using elastic nets, gradient boosted machines [GBMs] and shallow fully connected neural networks. We then hierarchically ensembled these models by eye dimension (Figure 1A and C).

We predicted chronological age with a testing R-Squared [R^2^] of 83.6±0.6% and a mean absolute error [MAE] of 2.62±0.05 years (Figure 2). The eye fundus images outperformed the OCT images as age predictors (R^2^=76.6±1.3% vs. 70.8±1.2%). The scalar-based models underperformed compared to the image-based model (R^2^=35.9±0.5). Between the different algorithms trained on all scalar features, the non-linear models outperformed the linear model (GBM: R^2^=35.8±0.6%; Neural network R^2^=30.8±2.4%; Elastic net: R^2^=25.0±0.4%).

**Figure 2:**
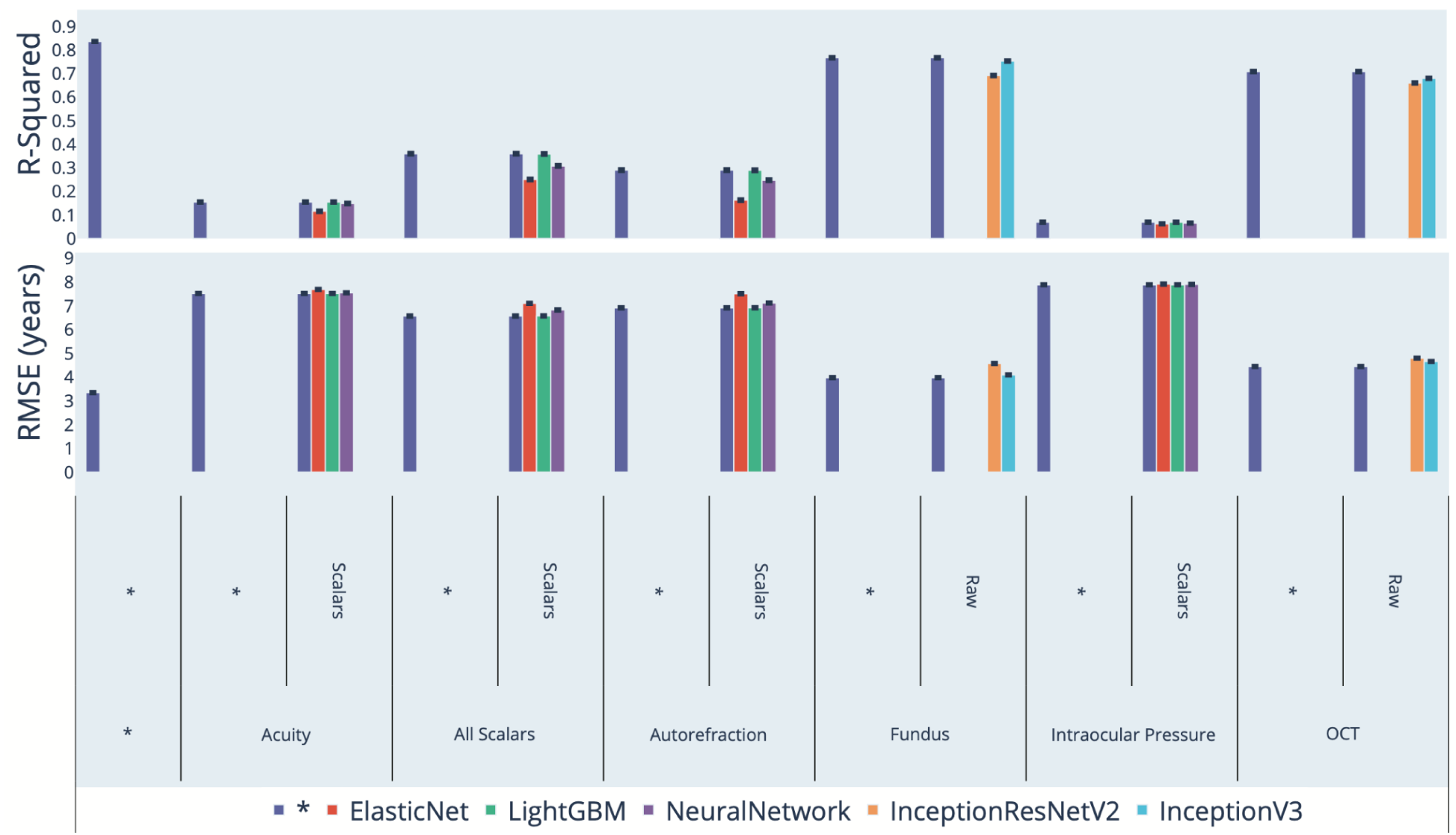
Chronological age prediction performance (R^2^ and RMSE) * represent ensemble models

We defined eye age as the prediction outputted by one of the models, after correction for the analytical bias in the residuals (Supplementary Methods and Methods). For example fundus-based eye age is the prediction outputted by the ensemble model trained on fundus images. If not specified otherwise, eye age refers to the prediction outputted by the best performing, all encompassing ensemble model.

We examined the relationship between the fundus and OCT predictors, specifically querying whether the predictions were correlated (does fundus age predict OCT age?). Fundus image-based and OCT image-based accelerated eye aging are modestly .239±.005 correlated. For comparison purposes, the two convolutional neural networks architectures (InceptionV3 and InceptionResNetV2) trained on the exact same dataset yielded accelerated aging definitions that are .762±.001 correlated (fundus images) and .815±.002 correlated (OCT images). (Figure 3).

**Figure 3:**
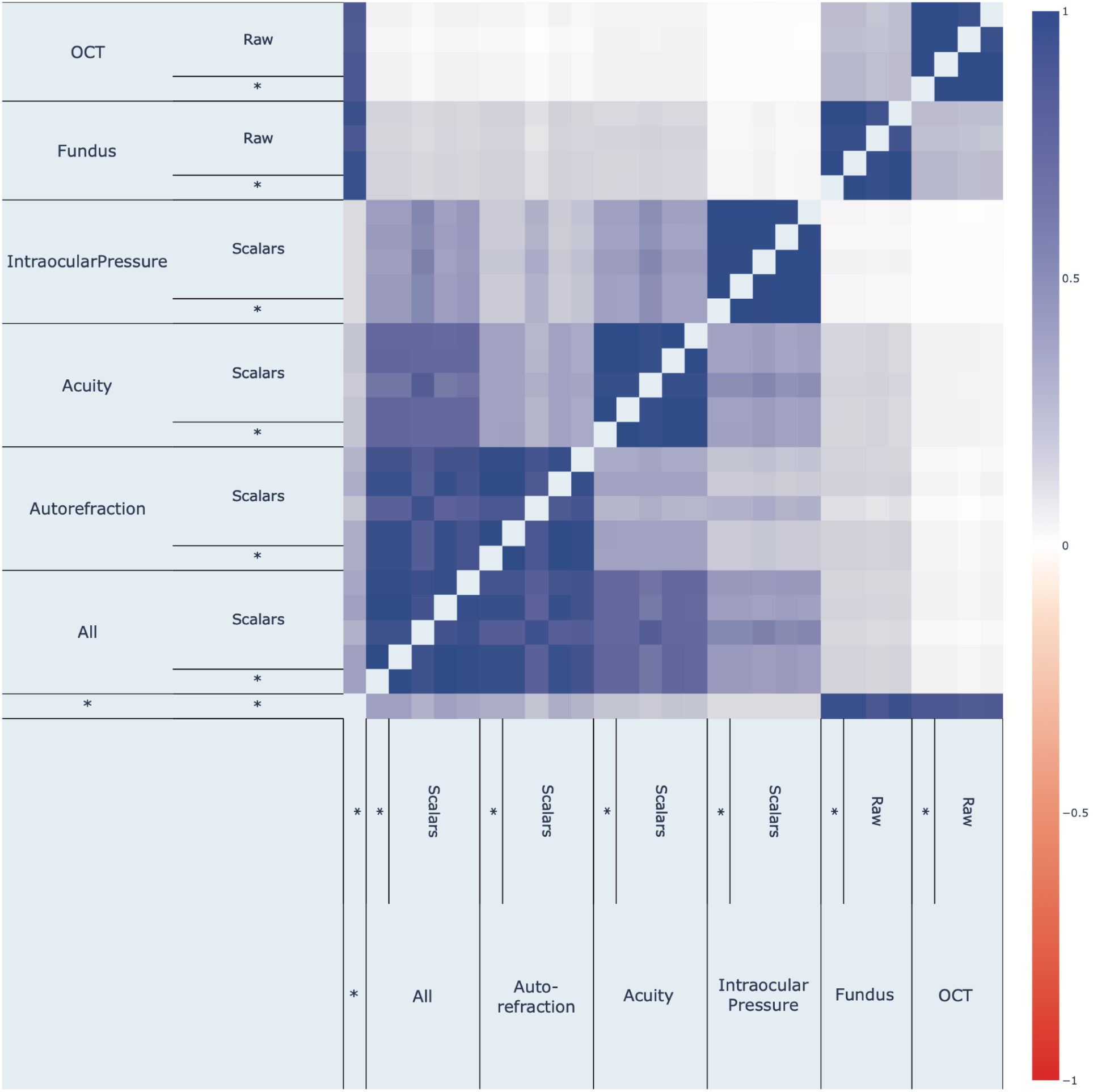
Phenotypic correlation between fundus-based and OCT-based accelerated eye aging.

### Genetic factors and heritability of accelerated eye aging

We performed genome wide association studies [GWASs] to estimate the GWAS-based heritability of general (h_g^2^=28.2±1.2%), fundus image-based (h_g^2^=26.0±0.9%), and OCT image-based (h_g^2^=23.6±0.9%) accelerated eye aging. We identified 26 (in proximity to 19 genes) and 42 genetic loci (212 genes) respectively the fundus and OCT “dimensions”, respectively. (Figure 4, Table S8; for quality control, see: Figure S12, Figure S13). Fundus image-based and OCT image-based accelerated eye aging are also genetically correlated (.299±.025).

**Figure 4:**
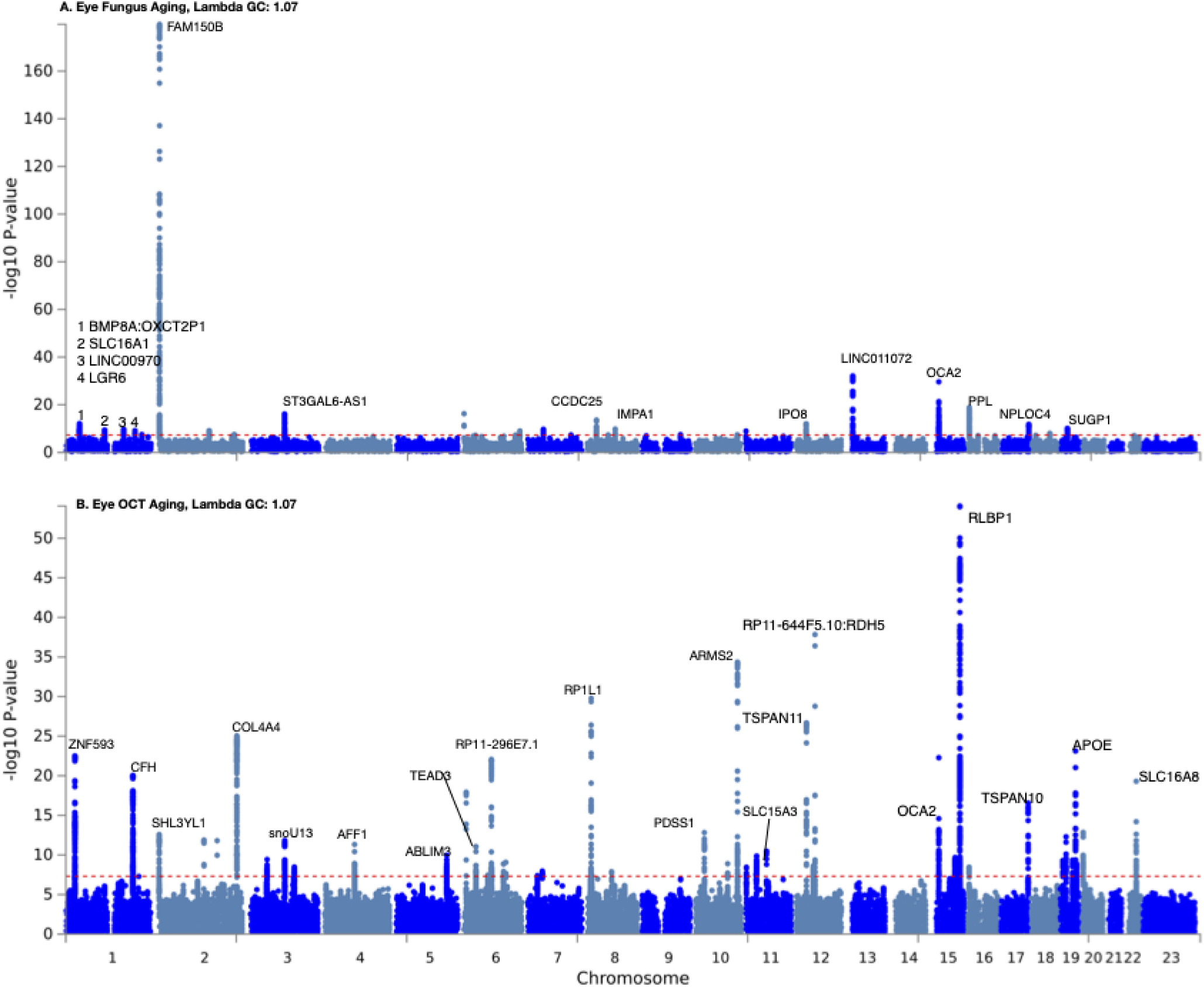
GWAS results for accelerated eye aging. **A.) Fundus image-based eye aging. B.) -OCT image-based eye aging.**-log10(p-value) vs. chromosomal position of locus. Dotted line denotes 5×10^-8^. Annotated genes are shown for independent loci whose minimum p-value are less than 1×10^-10^

The strongest signal associated with accelerated eye fundus aging included single nucleotide polymorphisms (SNPs, rs7605824, chromosome 2) in close proximity to *FAM150B*, (or *ALKAL2*), *ALK* and *LTK* ligand 2, (minimum p-value of all SNPs at locus: 2.4×10^-180^, Figure S14) and a cluster of genes that includes *SH3YL1*. Interestingly, we found a nearby locus, rs55742348 (pvalue of 3×10^-13^), associated with OCT aging. Specifically, rs55742348 was closest to the *SH3YL1* gene (Figure S15).

Another top signal included SNPs near *OCA2* (minimum p-value of 1×10^-30^), a gene that encodes the *P* protein involved in tyrosine transport for eventual melanin synthesis, or pigmentation of the eye. Interestingly, this locus was found in both eye fundus and OCT-aging predictions (minimum pvalue in OCT-based aging was 5×10^-23^). Other genes implicated included *SLC16A1*, *PPL, LGR6, NPLOC4,* and *SUGP1* (Figure 3A, and Table S8).

We found independent genetic loci associated with accelerated OCT-based aging that were different from those found in fundus-based eye aging (Figure 3B vs. Figure 3A) and specific to eye physiology. Briefly, these included a locus near *RLBP1* (retinaldehyde binding protein 1), a protein involved in vision and connected to diseases of the retina (Sparkes et al., 1992). We also identified variants near *ARMS2* (age-related maculopathy susceptibility 2), a protein a component of the choroidal (or the tissue between the retina and the inside of the eye). *RPL1L1* or retina pigmentosa like 1, is considered to be a paralog of retina pigmentosa 1(Bowne et al., 2003). Accelerated eye aging and age-related macular degeneration [AMD] are genetically positively correlated, but we were not powered in the UK Biobank to discriminate the correlation from zero (Table S9).

### Biomarkers, clinical phenotypes, diseases, environmental and socioeconomic variables associated with accelerated eye aging

We found that 4,372 biomarkers classified in 21 subcategories (Table S10), 187 clinical phenotypes classified in 11 subcategories (Table S13), 2,073 diseases classified in 26 subcategories (Table S16), 92 family history variables (Table S19), 265 environmental variables classified in nine categories (Table S22), and 91 socioeconomic variables classified in five categories (Table S25) were associated (Bonferroni-corrected p-value threshold of 0·05) with accelerated fundus and OCT eye aging. Please refer to the supplementary tables (Table S11, Table S12, Table S14, Table S15, Table S17, Table S18, Table S23, Table S24, Table S26, Table S27) for a summary of non-genetic factors associated with general, fundus-based and OCT-based accelerated eye aging. The exhaustive results can be found in Table S28 and explored at https://www.multidimensionality-of-aging.net/xwas/univariate_associations. We also compared fundus-based and OCT-based accelerated aging phenotypes in terms of their associations with non-genetic variables to understand if X-variables associated with accelerated aging in one eye dimension are also associated with accelerated aging in the other; while the OCT and fundus aging phenotypes have many non-shared genetic factors, they also, in contrast, share many environmental factor exposures associated with both (Figure S4). We discuss come of the specific findings below.

#### Biomarkers associated with accelerated eye aging

The three biomarker categories most associated with accelerated eye aging are urine biochemistry, blood pressure and eye intraocular pressure (Table S11). Specifically, 100.0% of urine biochemistry biomarkers are associated with accelerated eye aging, with the three largest associations being with microalbumin (correlation=.032), sodium (correlation=.027), and creatinine (correlation=.021). 100.0% of blood pressure biomarkers are associated with accelerated eye aging, with the three largest associations being with pulse rate (correlation=.075), systolic blood pressure (correlation=.044), and diastolic blood pressure (correlation=.040). 75.0% of eye intraocular pressure biomarkers are associated with accelerated eye aging, with the three largest associations being with right eye Goldmann-correlated intraocular pressure (correlation=.093), right eye corneal-compensated intraocular pressure (correlation=.092), and left eye Goldmann-correlated intraocular pressure (correlation=.090).

Conversely, the three biomarker categories most associated with decelerated eye aging are hand grip strength, spirometry and body impedance (Table S12). Specifically, 100.0% of hand grip strength biomarkers are associated with decelerated eye aging, with the two associations being with right and left hand grip strengths (respective correlations of .060 and .059). 100% of spirometry biomarkers are associated with decelerated eye aging, with the three associations being with forced expiratory volume in one second (correlation=.064), forced vital capacity (correlation=.060), and peak expiratory flow (correlation=.041). 100.0% of body impedance biomarkers are associated with decelerated eye aging, with the three largest associations being with whole body impedance (correlation=.040), left leg impedance (correlation=.038), and left arm impedance (correlation=.035).

#### Diseases associated with accelerated eye aging

The three disease categories most associated with accelerated eye aging are general health, cardiovascular diseases and eye disorders (Table S17). Specifically, 13.1% of general health variables are associated with accelerated eye aging, with the three largest associations being with personal history of disease (correlation=.039), problems related to lifestyle (correlation=.032), and presence of functional implants (correlation=.032). 11.7% of cardiovascular diseases are associated with accelerated eye aging, with the three largest associations being with hypertension (correlation=.061), chronic ischaemic heart disease (correlation=.030), and heart failure (correlation=.025). 11.4% of eye disorders are associated with accelerated eye aging, with the three largest associations being with cataract (correlation=.058),retinal disorders (correlation=.050), and retinal detachments and breaks (correlation=.041).

#### Environmental and behaviorial variables associated with accelerated eye aging

We define environmental variables as factors that are not assayed on the GWAS array and are potentially modifiable. 57.1% of variables that measure sleep are associated with accelerated eye aging, with the three largest associations being with napping during the day (correlation=.027), chronotype (being an evening person) (correlation=.026), and sleeplessness/insomnia (correlation=.026). 37.5% of smoking variables are associated with accelerated eye aging, with the three largest associations being with pack years adult smoking as proposition of lifespan exposed to smoking (correlation=.092), pack years of smoking (correlation=.090), and past tobacco smoking: smoked on most or all days (correlation=.066). 11.4% of physical activity variables are associated with accelerated eye aging, with the three largest associations being with time spent watching television (correlation=.063), no physical activity during the last four weeks among the ones listed in the questionnaire (correlation=.051), and types of transport used, work excluded: public transport (correlation=.042).

Conversely, the three environmental variable categories most associated with decelerated eye aging are physical activity, smoking and sleep (Table S24). Specifically, 57.1% of physical activity variables are associated with decelerated eye aging, with the three largest associations being with usual walking pace (correlation=.053), frequency of heavy do-it-yourself [DIY] work in the last four weeks (correlation=.046), and duration of heavy DIY (correlation=.044). 29.2% of smoking variables are associated with decelerated eye aging, with the three largest associations being with smoking status: never (correlation=.058), age started smoking (correlation=.057), and time from waking to first cigarette (correlation=.049). 28.6% of sleep variables are associated with decelerated eye aging, with the two associations being with snoring (correlation=.038) and getting up in the morning (correlation=.029).

### Identification of image and phenotypic features that drive eye age prediction

We give examples of the attention maps and a discussion of them in the Supplementary Results. We found that phenotypic measures of autorefraction, acuity, and intraocular pressure predicted age (Supplementary Results).

We also used methods for highlighting regions of the image that are driving the deep learning predictions. For fundus images, we found that GRAD-CAM attention maps highlighted regions of the eye that were highly vascularized (Figure S2), and the saliency maps highlighted the center of the eye (Supplementary Results). For the OCT images, all of the layers were highlighted and differed according to chronological age (Figure S3).

## Discussion

We built what is, to the best of our knowledge, the most accurate eye age predictor to date, and the comprehensive genetic, phenotypic, and environmental associations with them. We demonstrate that eye phenotype measures (fundus and OCT images, acuity, refractive error and intraocular pressure biomarkers) on which the predictor was trained capture different facets of eye aging, as demonstrated by (1) limited phenotypic correlation of the accelerated eye aging definitions that we derived from them, and (2) combining them into an ensemble model improved our age prediction accuracy (R^2^) by 7% compared to the best performing model built on a single dataset (R^2^=83·6±0·6% vs. 76·6±1·3%).

We identified genetic variants associated with accelerated eye aging, reflecting common, but modest, shared genetic architecture between fundus and OCT-based aging. We found many loci in genes whose biological function is thought to be localized to the eye, including *RP1L1* (RP1 Like 1)*, RLBP1* (Retinaldehyde-binding protein 1), and *OCA2* (melanocyte-specific transporter protein). Our GWAS signals included loci in *CFH* (complement factor H), and *ARMS2* (Age-Related Maculopathy Susceptibility 2). Further, we identified variants (p < 1×10^-23^) near *APOE* (Apolipoprotein E), a gene strongly connected to such as Alzheimer’s Disease (Farrer et al., 1997). We believe that these associations add to evidence supporting the biological relevance of our predictor.

Of particular interest includes a strong genetic signal on chromosome 2 near *FAM150B* (Figure S14), but also close to *SH3YL1* and *FAM110C* (p = 2·3×10^-180^). *FAM150B* is an “activating ligand” (along with *FAM150A*) to a neuronal receptor tyrosine kinase, *ALK(Guan et al., 2015)*, primarily investigated in its role in cancer (Reshetnyak et al., 2015), and recently, to treat persistent pain(Defaye et al., 2022) . We also find a locus with an association with OCT-aging close to *SH3YL1* (Figure S15, p = 3×10^-13^). To note, Ahadi and colleagues also find the same variants in fundus-based eye aging and have investigated their biological role in a model organism (personal communication).

Our genetic findings stand in contrast to age-related macular degeneration (AMD), which has a larger estimated GWAS-array based heritability (∼46% (Fritsche et al., 2015)), implying an altogether different genetic etiology of AMD vs. eye fundus/OCT aging. To note, our GWAS-based heritability estimates are smaller (∼28%) and the genetic associations that emerge implicate differing arrays of genes. However, there are notable similarities, such as *CFH* and *ARMS2,* which have been implicated in AMD (Fritsche et al., 2015; Vavvas et al., 2018).

Accelerated eye aging is also associated with biomarkers, phenotypes and diseases in other organ systems such as the cardiovascular system (e.g blood pressure, arterial stiffness, heart function, chest pain, hypertension, heart disease), metabolic health (e.g blood biochemistry, diabetes, obesity), brain health (e.g brain MRI biomarkers, mental health), the musculoskeletal system (e.g hand grip strength, heel bone densitometry, claudication, arthritis), mouth health, hearing, and others. Interestingly, accelerated eye aging is associated with facial aging. For example, diabetes increases the risk of vision loss or blindness (diabetic retinopathy (Duh et al., 2017)). Likewise, associations between cardiovascular and ocular health have been reported (De La Cruz et al., 2021). More generally, we found that accelerated eye aging is associated with a poor general health (e.g overall health rating, personal history of disease and medical treatment). We explore the connection between eye aging and other organ systems’ aging in a different paper (Le Goallec et al., 2021).

Similarly, accelerated eye aging is associated with eyesight biomarkers, clinical phenotypes and diseases (e.g wearing glasses/contacts, presbyopia, cataract, retinal disorders, retinal detachments and breaks).

In terms of environmental exposures, we found that general health factors such as poor sleep, smoking (including maternal smoking at birth) and lack of physical activity are associated with accelerated eye aging. Some diet variables such as cereal intake are associated with decelerated eye aging. Alcohol intake had a mixed association, with beer, cider and spirits being associated with accelerated eye aging, while usually taking alcohol with meals was associated with decelerated eye aging. We also found that playing video games and the time spent watching television were both associated with accelerated eye aging, which is coherent with the fact that screen time can strain the eye (computer vision syndrome) (Gowrisankaran and Sheedy, 2015). Associations between sleep (Dhillon et al., 2007), smoking (Galor and Lee, 2011), physical activity (Ong et al., 2018), alcohol intake (Kim, 2000), diet (Ohia et al., 2014) and ocular health have been reported previously. Participants with a higher socioeconomic status (e.g social support, income, education) were more likely to be decelerated eye agers, an effect likely due to an overall slower aging rate.

We used attention maps and scalar feature importances to identify the anatomical features driving eye age prediction. Our attention maps highlighted the eye fundus vascular features, which are consistent with Poplin et al.’s attention maps (Poplin et al., n.d.). Ege et al. reported that with age, eye fundus images tend to become more yellow because of optimal imperfections in the refractive media (Ege et al., 2002), which was possibly leveraged by our neural networks and could explain why some non-vascularized regions were also highlighted. Finally, our attention maps highlighted most retinal layers in the OCT images. The diversity of the features highlighted by the different models is coherent with the fact that age-related changes occur in all ocular tissues (Grossniklaus et al., 2013). The fact that eye age prediction is linked to spherical power and that the elastic net’s regression coefficient for this feature is positive is coherent with the fact that, with age, presbyopia becomes more prevalent (Glasser et al., 2001). Presbyopia –also called age-related farsightedness-is a consequence of the loss of the ability for the eye to accommodate to focus on nearby objects and it corresponds to positive spherical power. Similarly, cylindrical power is a measure of astigmatism and increases with age (Asano et al., 2005; Hayashi et al., 1993; Ho et al., 2010; Kim et al., 2002; Sim et al., 2017; Ueno et al., 2014), which explains its selection by the GBM and the positive regression coefficient in the elastic net.

We compare our models to the ones that can be found in the literature (Table S29). In terms of eye fundus images, chronological age was predicted from UKB samples with a R^2^ value of 74±1% and a MAE of 3·26 years by Poplin et al. (Poplin et al., 2018). We found that our ensemble model outperformed this prediction accuracy (R^2^=76·6±0·2; MAE=3·08±0·09 years). One potential explanation for this difference is that Poplin et al.’s model was trained on only 48,000 UKB participants, whereas our model was trained on 90,000 participants. Poplin et al.’s model was however also trained on 236,000 EyePACS (EyePACS, 2018) images, so it is unclear if the sample size benefited their model or ours and/or from the different CNN architectures. For example, our InceptionV3 model slightly outperformed their model (R^2^=75·2±0·1%), whereas our InceptionResNetV2 model significantly underperformed (R^2^=69·1±0·2%). More recently Nagasato et al. used transfer learning to train a VGG16 architecture on a far smaller sample size (n=85) using ultra-wide-field pseudo-color eye fundus images collected from participants aged 57·5±20·9 years to predict chronological age. However, they did not report their R^2^ value, only the slope of the regression coefficient between their predictor and chronological age (standard regression coefficient=0·833) (Nagasato et al., 2020). In terms of optical coherence tomography images, Chueh et al. predicted age by using transfer learning and training a ResNet18 architecture on 6,147 OCT images from 20-90 year-old participants (MAE=5·78±0.29) (Chueh et al., n.d.). We outperformed this model (MAE=3·49±0·07 years), perhaps due to our greater sample size (N=173,695).

Other researchers leveraged eye features not available in the UK Biobank. Sgroi et al., Erbilek et al. and Rajput and Sable (Erbilek et al., 2013; Rajput and Sable, n.d.; Sgroi et al., 2013) predicted chronological age from iris images. Sgroi et al. trained a random forest on 630 scalar texture features extracted from 596 sample iris images to classify participants aged 22-25 years versus participants older than 35 years with an accuracy of 64%. Erbilek et al. built an ensemble of five simple geometric features extracted from the same iris images dataset. Using what they refer to as the sensitivity negotiation method, inspired by game theory, they obtained a classification accuracy of 75%. Rajput and Sable trained CNNs on 2,130 iris images collected from 213 participants aged 3-74 years and predicted chronological age with an MAE of 5-7 years (Rajput and Sable, n.d.). Bobrov et al. trained CNNs on 8,414 eye corner images collected from participants aged 20-80 and predicted chronological age with a R^2^ value of 90·25% (estimated from the Pearson correlation) and a MAE of 2·3 years (Bobrov et al., 2018).

We identify three limitations to our study. First, we extracted a 2D image from each 3D OCT image to limit the need for computational resources. Leveraging the full data with a three-dimensional convolutional neural network architecture may increase the age prediction accuracy. The GWAS and environmental factor correlations that we report do not allow us to infer causality. Third, we did not distinguish between left eye and right eye aging. A possible future direction would be to test if, on average, the right and left eyes age at significantly different rates.

In conclusion, our eye age predictor can be used to monitor the eye aging process. The genetic and non-genetic associations we found also suggest potential lifestyle and therapeutic interventions to slow or reverse eye aging. The GWAS in particular could shed light on the etiology of age-related eye disorders such as age-related macular degeneration (Winkler et al., 2020). Finally, our eye age predictor could be used to evaluate the effectiveness of emerging rejuvenating therapies (de Magalhães et al., 2017) on the eye. The DNA methylation clock is for example already used in clinical trials (Duke Clinical Research Institute, Elysium Health, 2019; Horvath, 2013; Horvath et al., 2014), but as aging is multidimensional (Le Goallec et al., 2021; Li et al., 2020), multiple clocks might be needed to properly assess the effect of a rejuvenating therapeutic intervention on the different portions of the eye.

## Data Availability

We used the UK Biobank (project ID: 52887). The code can be found at https://github.com/Deep-Learning-and-Aging. The results can be interactively and extensively explored at https://www.multidimensionality-of-aging.net/. We will make the biological age phenotypes available through UK Biobank upon publication. The GWAS results can be found at https://www.dropbox.com/s/59e9ojl3wu8qie9/Multidimensionality_of_aging-GWAS_results.zip?dl=0.

https://www.multidimensionality-of-aging.net/

## Author Contributions

All authors had access to the data during the project.

**Alan Le Goallec:** (1) Designed the project. (2) Supervised the project. (3) Predicted chronological age from images. (4) Computed the attention maps for the images. (5) Ensembled the models, evaluated their performance, computed biological ages and estimated the correlation structure between the eye aging dimensions. (6) Performed the genome wide association studies. (5) Designed the website. (6) Wrote the manuscript.

**Samuel Diai:** (1) Predicted chronological age from scalar features. (2) Coded the algorithm to obtain balanced data folds across the different datasets. (3) Wrote the python class to build an ensemble model using a cross-validated elastic net. (4) Performed the X-wide association study. (5) Implemented a first version of the website https://www.multidimensionality-of-aging.net/.

**Sasha Collin:** (1) Preprocessed the OCT images. (2) Preprocessed the fundus images.

**Théo Vincent:** (1) Website data engineer. (2) Implemented a second version of the website https://www.multidimensionality-of-aging.net/.

**Chirag J. Patel:** (1) Designed the project. (2) Supervised the project. (2) Wrote the manuscript. (3) Performed analyses, (4) Provided funding.

## Acknowledgments

We would like to thank Raffaele Potami from Harvard Medical School research computing group for helping us utilize O2’s computing resources. We thank HMS RC for computing support. We also want to acknowledge UK Biobank for providing us with access to the data they collected. The UK Biobank project number is 52887.

## Conflicts of Interest

None.

## Funding

NIEHS R01ES032470

NIAID R01AI127250

MassCATS, Massachusetts Life Science Center

Sanofi

The funders had no role in the study design or drafting of the manuscript(s).

## Supplementary Appendix

## Results

### Biomarkers, clinical phenotypes, diseases, environmental and socioeconomic variables associated with accelerated eye aging

Below, we summarize the findings for accelerated general eye aging.

#### Clinical phenotypes associated with accelerated eye aging

The three clinical phenotype categories most associated with accelerated eye aging are chest pain, breathing and general health (Table S14). Specifically, 100.0% of chest pain phenotypes are associated with accelerated eye aging, with the three largest associations being with chest pain or discomfort walking normally (correlation=.033), chest pain due to walking ceases when standing still (correlation=.030), and chest pain or discomfort (correlation=.030). 100.0% of breathing phenotypes are associated with accelerated eye aging, with the two associations being with shortness of breath walking on level ground (correlation=.059) and wheeze or whistling in the chest in the last year (correlation=.051). 62.5% of general health phenotypes are associated with accelerated eye aging, with the three largest associations being with overall health rating (correlation=.085), long-standing illness, disability or infirmity (correlation=.071), and falls in the last year (correlation=.032).

Conversely, the three clinical phenotype categories most associated with decelerated eye aging are sexual factors (age first had sexual intercourse: correlation=.030), mouth health (no mouth/teeth problems: correlation=.019) and general health (no weight change during the last year: correlation=.042). (Table S15)

The “diseases” associated with decelerated eye aging are related to pregnancy and delivery (perineal laceration during delivery: correlation=.029; single spontaneous delivery: correlation=.019; outcome of delivery: correlation=.044; supervision of high-risk pregnancy: correlation=.020). (Table S18)

#### Socioeconomic variables associated with accelerated eye aging

The three socioeconomic variable categories most associated with accelerated eye aging are sociodemographics (private healthcare: correlation=.020), social support (leisure/social activities: none of the listed: correlation=.021) and household (renting from local authority, local council or housing association: correlation=.043; type of accommodation: flat, maisonette or apartment: correlation=.032). (Table S26)

Conversely, the three socioeconomic variable categories most associated with decelerated eye aging are social support, sociodemographics and household (Table S27). Specifically, 22.2% of social support variables are associated with decelerated eye aging, with the two associations being leisure/social activities: sports club or gym (correlation=.026) and being able to confide (correlation=.019). 14.3% of sociodemographic variables are associated with decelerated eye aging (one association: not receiving attendance/disability/mobility allowance. correlation=.048). 11.4% of household variables are associated with decelerated eye aging, with the three largest associations being with number of people in the household (correlation=.057), average total household income before tax (correlation=.054), and number of vehicles (correlation=.048).

### Identification of image and phenotypic features that drive eye age prediction

We best predicted eye aging with a GBM (R^2^=35.8±0.6%) trained on autorefraction (35 features), acuity (eight features) and intraocular pressure (eight features) measures, along with sex and ethnicity. Autorefraction measures alone predicted age with a R^2^ of 29.0±0.2%, acuity measures with a R^2^ of 15.5±0.2% and intraocular pressure with a R^2^ of 6.9±0.1%. Specifically, the associated most important scalar features for the model including all the predictors are (1; 2) the spherical power (right and left eyes), (3; 4) the cylindrical power (right and left eyes), (5; 6) the astigmatism (right and left eyes), (7) the 3mm asymmetry index (left eye), (8) the 6mm cylindrical power (right eye), (9) the 3mm asymmetry angle (left eye) and (10) the 6mm weak meridian angle. In the linear context of the elastic net (R^2^=25.0±0.4%), the spherical power, the cylindrical power, the 6mm cylindrical power and the 6mm weak meridian angle are associated with older age, whereas the astigmatism angle, the 3mm asymmetry index and the 3mm asymmetry angle are associated with younger age. The feature importances for all scalar-based models can be found in Table S2, Table S3, Table S4 and Table S5.

In terms of eye fundus images, the Grad-RAM attention maps tended to highlight the eye regions that were highly vascularized, whereas the saliency maps also frequently highlighted the center of the eye (Figure S2). In terms of OCT images, Grad-RAM highlighted all the retinal layers with different regions emphasized for different participants. Similarly, saliency maps highlighted different layers for different participants, as well as the fovea. The resolution of the resized images makes it difficult to precisely identify these layers, but the list seems to include the posterior hyaloid surface, the inner limiting membrane, the external limiting membrane, ellipsoid portion of the inner segments, the cones outer segment tips line and the retinal pigment epithelium (Figure S3).

### Correlation between fundus image-based and OCT image-based accelerated eye aging

We compared fundus-based and OCT-based accelerated aging phenotypes in terms of their associations with non-genetic variables to understand if X-variables associated with accelerated aging in one eye dimension are also associated with accelerated aging in the other. For example, in terms of environmental variables, are the diets that protect against eye aging in terms of fundus image the same as the diets that protect against eye aging in terms of OCT image? We found that the correlation between accelerated brain anatomical and cognitive aging is .926 in terms of biomarkers, .704 in terms of associated clinical phenotypes, .830 in terms of diseases, .838 in terms of environmental variables and .748 in terms of socioeconomic variables (Figure S4). These results can be interactively explored at https://www.multidimensionality-of-aging.net/correlation_between_aging_dimensions/xwas_univariate.

## Methods

### Hardware

We performed the computation for this project on Harvard Medical School’s compute cluster, with access to both central processing units [CPUs] and general processing units [GPUs] (Tesla-M40, Tesla-K80, Tesla-V100) via a Simple Linux Utility for Resource Management [SLURM] scheduler.

### Software

We coded the project in Python (Van Rossum and Drake, 2011) and used the following libraries: NumPy (Oliphant, 2006; Walt et al., 2011), Pandas (McKinney and Others, 2010), Matplotlib (Hunter, 2007), Plotly (Inc, 2015), Python Imaging Library (Clark, 2018), SciPy (Millman et al., 2011; Oliphant, 2007; Virtanen et al., 2020), Scikit-learn (Pedregosa et al., 2011), LightGBM (Ke et al., 2017), XGBoost (Chen and Guestrin, 2016), Hyperopt (Bergstra et al., 2013b), TensorFlow 2 (Abadi et al., 2015), Keras (Chollet and Others, 2015), Keras-vis (Kotikalapudi and Others, 2019), iNNvestigate (Alber et al., 2019). We used Dash (Hossain et al., 2019) to code the website on which we shared the results. We set the seed for the os library, the numpy library, the random library and the tensorflow library to zero.

### Data Preprocessing

#### Demographic variables

First, we removed out the UKB samples for which age or sex was missing. For sex, we used the genetic sex when available, and the self-reported sex when genetic sex was not available. We computed age as the difference between the date when the participant attended the assessment center and the year and month of birth of the participant to estimate the participant’s age with greater precision. We one-hot encoded ethnicity.

#### Scalar biomarkers: acuity, autorefraction and intraocular pressure

We define scalar data as a variable that is encoded as a single number, such as eye spherical power or astigmatism angle, as opposed to data with a higher number of dimensions, such as images. The complete list of scalar biomarkers can be found in Table S10 under “Eye”. We did not preprocess the scalar data, aside from the normalization that is described under cross-validation further below.

#### Images

##### Eye fundus

The UKB contains eye fundus RGB images, with dimensions 1536*2048*3 pixels. Data from both the left eye (field 21015, 87,562 samples for 84,760 participants were collected) and the right eye (field 21016, 88,259 samples for 85,239 participants) was collected. We took the vertical symmetry of left eye images. We cropped the images to remove the black border surrounding the actual eye fundus images, which yielded centered square images with dimensions 1388*1388*3. Some of the images were low quality on visual inspection. For example, they had very low or very high luminosity. To reduce the prevalence of such images, we developed the following two-step heuristic. First, we computed the mode of the distribution for each of the three RGB layers. If the mode is strictly larger than 250 for both the red, the green and the blue channels, and has a frequency at least as high as 100,000 for the three channels, the image is removed. Second, we compute the median of the distribution for each of the three RGB channels. If the difference between the red median and the maximum between the green and blue medians is strictly smaller than ten, the image is removed. A sample of preprocessing OCT images can be found in Figure S5.

##### Eyes optical coherence tomography

The UKB contains (optical coherence tomography) OCT 3D images from both the left eye (field 21017, 87,595 samples for 84,173 participants) and the right eye (field 21018, 88,282 images for 85,262 participants) (Figure S6). Each sample contains 128 images of dimensions 650*512*1 grayscale pixels. We selected the image where the fovea pit is the deepest as the input for our models, as follows. First, for each sample, we discarded images before the 30th image and after the 105th image, because the fovea was never the deepest in the first or last images. Then we applied the fastNlMeansDenoising function from the openCV python library (BRADSKI and G, 2000) to each of the remaining 75 images with the following hyperparameter values: h=30, templateWindowSize=7, searchWindowSize=7. We then applied OpenCV’s Canny edge algorithm (Canny, 1986) with 30 and 150 as the two thresholds for the hysteresis procedure to detect the surface of the retina. Instead of relying on a single threshold to detect edges, the hysteresis method consists in setting both an upper and lower threshold. Pixels whose gradient value is above the upper threshold are classified as belonging to an edge, and pixels whose gradient value is below the lower threshold are classified as non-edge. The remaining pixels, whose gradient value is between the two thresholds, are classified as an edge if they are connected to at least one edge pixel. For each column of pixels, we identified the surface of the retina as the first pixel encountered set to 1 by the canny edge algorithm, starting from the top of the image. We applied two smoothing methods (triangle moving average and Savitzky–Golay filter) to detect the surface (Figure S7).

We then computed the curvature of the detected surface (Figure S8), and we used the maximum curvature value for each of the 75 images. Next, we identified which of these 75 images had the maximal curvature, corresponding to the pit of the fovea. The list of maximal curvature for each image is a noisy function, so we applied the Savitzky–Golay filter to identify the image with the maximal curvature (Figure S9). Finally, we cropped the selected image to center the fovea pit vertically to obtain the final images with dimension 500*512*3 pixels (Figure S10). A sample of preprocessed OCT images can be found in Figure S11.

To resolve prohibitory long training times, we resized the images so that the total number of pixels for each channel would be below 100,000.

#### Data augmentation

To prevent overfitting and increase our sample size during the training we used data augmentation (Shorten and Khoshgoftaar, 2019) on the images. Each image was randomly shifted vertically and horizontally, as well as rotated. We chose the hyperparameters for these transformations’ distributions to represent the variations we observed between the images in the initial dataset. A summary of the hyperparameter values for the transformations’ distributions can be found in Table S31.

The data augmentation process is dynamically performed during the training. Augmented images are not generated in advance. Instead, each image is randomly augmented before being fed to the neural network for each epoch during the training.

### Machine Learning Algorithms

#### Scalar data

We used three different algorithms to predict age from scalar data (non-dimensional variables, such as laboratory values). Elastic Nets [EN] (a regularized linear regression that represents a compromise between ridge regularization and LASSO regularization), Gradient Boosted Machines [GBM] (LightGBM implementation (Ke et al., 2017)), and Neural Networks [NN]. The choice of these three algorithms represents a compromise between interpretability and performance. Linear regressions and their regularized forms (LASSO (Tibshirani, 1996), ridge (Hoerl and Kennard, 1970), elastic net (Zou and Hastie, 2005)) are highly interpretable using the regression coefficients but are poorly suited to leverage non-linear relationships or interactions between the features and therefore tend to underperform compared to the other algorithms. In contrast, neural networks (Popescu et al., 2009; Rosenblatt, 1958) are complex models, which are designed to capture non-linear relationships and interactions between the variables. However, tools to interpret them are limited (Ribeiro et al., 2016) so they are closer to a “black box”. Tree-based methods such as random forests (Breiman, 2001), gradient boosted machines (Friedman, 2001) or XGBoost (Chen and Guestrin, 2016) represent a compromise between linear regressions and neural networks in terms of interpretability. They tend to perform similarly to neural networks when limited data is available, and the feature importances can still be used to identify which predictors played an important role in generating the predictions. However, unlike linear regression, feature importances are always non-negative values, so one cannot interpret whether a predictor is associated with older or younger age. We also performed preliminary analyses with other tree-based algorithms, such as random forests (Breiman, 2001), vanilla gradient boosted machines (Friedman, 2001) and XGBoost (Chen and Guestrin, 2016). We found that they performed similarly to LightGBM, so we only used this last algorithm as a representative for tree-based algorithms in our final calculations.

#### Images

##### Convolutional Neural Networks Architectures

We used transfer learning (Pan and Yang, 2010; Tan et al., 2018; Weiss et al., 2016) to leverage two different convolutional neural networks (LeCun et al., 2015) [CNN] architectures pre-trained on the ImageNet dataset (Deng et al., 2009; Krizhevsky et al., 2012; Russakovsky et al., 2015) and made available through the python Keras library (Chollet and Others, 2015): InceptionV3 (Szegedy et al., 2016) and InceptionResNetV2 (Szegedy et al., 2017). We considered other architectures such as VGG16 (Simonyan and Zisserman, 2014), VGG19 (Simonyan and Zisserman, 2014) and EfficientNetB7 (Tan and Le, 2019), but found that they performed poorly and inconsistently on our datasets during our preliminary analysis and we therefore did not train them in the final pipeline. For each architecture, we removed the top layers initially used to predict the 1,000 different ImageNet images categories. We refer to this truncated model as the “base CNN architecture”.

We added to the base CNN architecture what we refer to as a “side neural network”. A side neural network is a single fully connected layer of 16 nodes, taking the sex and the ethnicity variables of the participant as input. The output of this small side neural network was concatenated to the output of the base CNN architecture described above. This architecture allowed the model to consider the features extracted by the base CNN architecture in the context of the sex and ethnicity variables. For example, the presence of the same anatomical feature can be interpreted by the algorithm differently for a male and for a female. We added several sequential fully connected dense layers after the concatenation of the outputs of the CNN architecture and the side neural architecture. The number and size of these layers were set as hyperparameters. We used ReLU (Agarap, 2018) as the activation function for the dense layers we added, and we regularized them with a combination of weight decay (Bos and Chug, n.d.; Krogh and Hertz, 1992) and dropout (Srivastava et al., 2014), both of which were also set as hyperparameters. Finally, we added a dense layer with a single node and linear activation to predict age.

##### Compiler

The compiler uses gradient descent (Bottou et al., 2018; Ruder, 2016) to train the model. We treated the gradient descent optimizer, the initial learning rate and the batch size as hyperparameters. We used mean squared error [MSE] as the loss function, root mean squared error [RMSE] as the metric and we clipped the norm of the gradient so that it could not be higher than 1.0 (Zhang et al., 2019).

We defined an epoch to be 32,768 images. If the training loss did not decrease for seven consecutive epochs, the learning rate was divided by two. This is theoretically redundant with the features of optimizers such as Adam, but we found that enforcing this manual decrease of the learning rate was sometimes beneficial. During training, after each image has been seen once by the model, the order of the images is shuffled. At the end of each epoch, if the validation performance improved, the model’s weights were saved.

We defined convergence as the absence of improvement on the validation loss for 15 consecutive epochs. This strategy is called early stopping (Prechelt, 1998) and is a form of regularization. We requested the GPUs on the supercomputer for ten hours. If a model did not converge within this time and improved its performance at least once during the ten hours period, another GPU was later requested to reiterate the training, starting from the model’s last best weights.

### Training, tuning and predictions

#### Data splitting

We split the 676,787 samples into ten data folds, while keeping all samples from the same participant in the same fold. To ensure this, we split the 502,211 participants’ ids (referred to by UKB as “eid”) into ten different buckets of the same size. To generate ten folds for each sub-dataset (e.g. autorefraction biomarkers), we took the intersection of the samples in each of the ten folds with the samples for which the sub-dataset data was available. This method had however one important loophole, which is that we could not guarantee that the folds for the sub-datasets would be balanced. For example, autorefraction biomarkers were only recorded for 120,000 out of the 500,000 participants. Since the 500,000 participants are split into ten folds, a fold contains approximately 50,000 participants. Although unlikely, we could therefore not guarantee that all or most of the autorefraction samples would be attributed to the three first data folds, leading to highly unbalanced folds for the autorefraction analysis. Unbalanced folds can lead to problems during the cross-validation (see further below), as models trained on a smaller number of samples will tend to generalize worse. One solution would have been to use a different split for each dataset, but this would have generated problems when building the ensemble models fold by fold (see Methods - Models ensembling). To mitigate this issue of unbalanced data folds, we developed the following heuristic. We randomly split the 502,211 participants into ten folds, 1,000 times. For each of these 1,000 splits, we computed for each sub-dataset the variance of the percentages of samples in each fold. We then scored each of the 1,000 splits using the maximum of the variance among the different sub-datasets. For example, if the autorefraction samples were not evenly split for the ith split out of the 1,000 splits (e.g. fold 1: 55% of the samples, every other fold: 5% of the samples), the variance of the sample proportions would be high, which would yield a poor score for the ith split. Finally, we selected the split with the lowest score as the final split for the main dataset, and for all the sub-datasets. This selected split had a score of 5.8e-4, which means that the most unbalanced sub-dataset had a variance in its sample size proportion between its ten folds of 5.8e-4.

#### Scalar data

##### Nested cross-validation

Cross-validation is a method to tune the regularization of models and prevent overfitting (Kohavi and Others, 1995). For the models inputting scalar data (Figure 1A in green), we tuned the hyperparameters and generated a testing prediction for each sample using a nested 10×9-folds cross-validation. We refer to the two nested cross-validations as the “outer” and the “inner” cross-validations. The outer-cross validation is used to generate an unbiased testing prediction for each sample, as opposed to a simple split of the data into a “training+validation” set on one hand, and a testing set on the other hand, which would only generate a testing prediction for one tenth of the dataset. The inner cross-validation is used to tune the hyperparameters more precisely, leveraging the full inner cross-validation dataset as a validation set, as opposed to a simple data split of the “training+validation” dataset into a training and a validation sets, which would only use one data fold as the validation set to estimate the performance associated with a specific combination of hyperparameters. The nested cross-validation is illustrated in Table S33.

##### Bayesian hyperparameters optimization

To tune the hyperparameters, we used the Tree-structured Parzen Estimator Approach (Bergstra et al., 2011) [TPE] of the hyperopt python package (Bergstra et al., 2013a). TPE is a sequential Bayesian hyperparameters optimization method that iteratively suggests the next most promising hyperparameters combination as a function of the hyperparameters combinations that have already been tested, by building a probabilistic representation of the objective function. We set the number of iterations to 30. For each model, 30 different hyperparameter combinations are iteratively tested before selecting the best performing one. The hyperparameters names and their ranges defining the hyperparameters space can be found in Table S32. It might be of interest to other researchers that we initially tuned the hyperparameters using a random search (Bergstra and Bengio, 2012) with the same number of iterations, and we did not observe a significant improvement in the model’s performance after implementing the Bayesian hyperparameters optimization.

##### Example

For the sake of clarity, let us walk through a concrete example, which is illustrated in Table S33. Suppose we want to generate unbiased predictions for every sample in a dataset using an elastic net. First, let us generate the testing prediction for the data fold F9, which is performed by the first fold of the outer cross-validation (outer cross-validation fold 0). We select the data fold F9 out of the ten data folds as the testing fold, and we select the remaining nine data folds as “training+validation” folds for the inner cross-validation. We scale and center the target (age) and the predictors using the mean and standard deviation values of the variables on the “training+validation” dataset. We then enter the first inner-cross validation.

For the first inner cross-validation fold, we select the data fold F8 as the validation set, and the remaining eight “training+validation” data folds as the training set. We re-scale and center age and the predictors in the training and the validation sets using the mean and standard deviation values of the training set. We train the model on the eight training data folds with the first hyperparameters combination sampled by the TPE algorithm (one value for alpha and one value for l1_ratio) and generate validation predictions on the validation fold (data fold F8), which we unscale. This completes the first of the nine inner cross-validation folds (Inner CV fold 0). We then permute the nine inner data folds. We scale the age and the predictors using the mean and standard deviation computed on the new training set. Then we train the model with the same first combination of hyperparameters on eight data folds, leaving aside the data fold F9 (still being used as the testing set for the outer cross-validation) and the data fold F7 (now being used as the validation set for the inner cross-validation). We then use the new trained model to generate validation predictions on the data fold F7, which we unscale. This completes the second of the nine inner-cross validation folds (Inner CV fold 1). We then reiterate these inner permutation and training processes seven more times, until every data fold in the nine “training+validation” data folds is used as the validation set once. At this point, we concatenate the validation predictions from these nine validation folds to obtain the overall validation predictions associated with the first hyperparameters combination, and compute the associated performance metric (e.g. RMSE). This completes the inner-cross validation for the first hyperparameters combination.

We then perform the same 9-folds inner cross-validation, this time with the second hyperparameters combination suggested by the TPE algorithm. We iterate this process 28 more times, until 30 different hyperparameters combinations have iteratively been tested. Next, we select the hyperparameter combination that yielded the best validation performance (e.g. minimum RMSE), and we retrain a model on the whole nine “training+validation” data folds (all data folds except for data fold #1), using this best performing hyperparameters combination. This completes the first inner cross-validation.

We then use the model to generate unbiased predictions on the unseen testing set (data fold F9) and record these predictions. By anticipation for the ensembling algorithm (see Methods - Models ensembling) we also need to compute validation predictions on the data fold F8. We do this by training a model on all the data folds aside from the validation fold (data fold F8) and the testing fold (data fold F9), with the selected hyperparameters combination. We then use this trained model to compute predictions on the validation fold (data fold F8) and record these predictions, after unscaling them. This completes the first of the ten outer cross-validation folds (outer cross-validation 0).

We then complete the second outer cross-validation fold (outer cross-validation 1), this time using the data fold F8 as the testing dataset, to obtain unbiased testing predictions on this data fold, as well as validation predictions on the data fold F7. We reiterate the process eight more times to obtain the testing and validation predictions on the remaining data folds. We then concatenate the testing predictions from the ten data folds to obtain our final testing predictions for the model. Similarly, we concatenate the validation predictions from the ten data folds to obtain our final testing predictions for the model, which will later be used during ensemble models building and model selection (see Methods - Models ensembling).

The final validation and testing predictions for each data fold are therefore not necessarily associated with the same hyperparameters combination. It is also important to notice that we performed a single outer cross-validation, but that we performed a separate inner-cross validation for each outer cross-validation fold (hence the word “nested”), for a total of ten inner cross-validations per outer cross-validation fold.

#### Images

##### Hyperparameters tuning upstream of the cross-validation

The hyperparameters we tuned were the number of added fully connected dense layers, the number of nodes in these layers, their activation function, the optimizer, the initial learning rate, the weight decay, the dropout rate, the data augmentation amplitude and the batch size. Repeatedly tuning the values of the hyperparameters for different deep neural networks architectures and on the different cross-validation folds would have been prohibitively time and resource consuming. Instead, we sequentially explored how each hyperparameter was affecting the training and validation performances for a single architecture (InceptionV3) on a single cross validation fold (fold #0, see Methods - Training, tuning and predictions - Images - Cross-validation for the detailed description of the cross-validation). We then extrapolated the hyperparameter values to the other architectures, datasets and cross-validation folds. The hyperparameters combinations tested during the tuning can be found in Table S34.

First, we maximized the batch size for each architecture. The maximum number of images per batch depends on the memory of the GPU and the size of the architecture, which itself depends on the dimensions of the image. We used a batch size of 32 for InceptionV3 and 8 for InceptionResNetV2.

Then, we tested the learning rates, including 1e-6, 1e-5, 1e-4, 1e-3, 1e-2 and 1e-1. We observed that learning rates larger than 1e-4 prevented the model from converging for some runs. Second, we did not observe significant differences between the results obtained with learning rates smaller than 1e-4. We therefore set the initial learning rate to be 1e-4 for all models to shorten the time to convergence while ensuring that the learning rate was small enough to allow convergence and the finding of a local minima for the loss function.

Then we tested three different optimizers to perform the gradient descent: Adam (Kingma and Ba, 2014), Adadelta (Zeiler, 2012) and RMSprop (Hinton, n.d.). We did not observe any significant differences between the optimizers, so we set the optimizer to be Adam.

We then added different numbers of fully connected layers between the base CNN and side CNN’s concatenated outputs and the final activation layer. We set the number of nodes to be 1,024 in the first added layer and then decreased the number of nodes by a factor of two for each successive layer. For example, if we added three fully connected layers, the number of nodes was 1024, 512 and 256. We added zero, one and five layers. We did not observe significant differences in the performance of the different architectures, so we set the number of fully connected layers to one.

We then tested powers of two from 16 to 2,048 as the number of nodes in this single layer. We did not observe significant differences between these architectures, so we set the number of nodes to be 1,024 to keep the number close to the initial number of nodes in the imported CNN architectures, as these were initially used to perform classification between 1,000 categories.

We tested two different activation functions for the activation functions of the fully connected layers we added in the side neural network and before the final linear layer. We did not observe any significant differences between the rectified linear units [ReLU] (Nair and Hinton, 2010) and the scaled exponential linear units [SELU] (Klambauer et al., 2017) as activation functions, so we used the more common ReLU.

We then tested different levels of data augmentation. We introduced a hyperparameter that we called “data augmentation factor”. The data augmentation factor modulates the amount of variation introduced by the data augmentation, while preserving the ratio between the different transformations. For example, a data augmentation factor of one is equivalent to the default data augmentation (see Preprocessing - Data augmentation - Images), but a data augmentation factor of two will double the ranges of the possible values sampled and the expected values for the vertical shift, the horizontal shift, the rotation and the zoom on the original images. We tested the following values for the data augmentation factor: 0, 0.1, 0.5, 1, 1.5 and 2. We found that different values for the data augmentation factor hyperparameter yielded similar results, as long as the data augmentation factor was not zero. We therefore set the data augmentation factor to be one when training the final models.

We then tuned the dropout rate for the fully connected layers we added. We tested the following values: 0, 0.1, 0.25, 0.3, 0.5, 0.75, 0.9 and 0.95. We observed that a dropout rate of 0.95 led to underfitting and that smaller values reduced overfitting on the training set but without improving the validation performance. As a consequence, we used a dropout rate of 0.5.

Finally, we tuned the weight decay. We tested the following values: 0, 0.1, 0.2, 0.3, 0.4, 0.5, 1, 5, 10 and 100. For the larger datasets, we found that weight decay values as low as 0.4 could lead to underfitting. We found that lower weight decay values reduced overfitting on the training set without significantly improving the validation performance. We set the weight decay to 0.1.

Altogether, we found that hyperparameter tuning had little effect on the validation performance as long as extreme hyperparameters values were not selected.

##### Cross-validation

Training deep convolutional neural networks on images and videos is too time and resource consuming to perform a nested cross-validation. Therefore, we tuned the hyperparameters during the preliminary analysis, as described above. After hyperparameters tuning, we performed a simple outer cross-validation to obtain a testing prediction for each sample of the datasets, but we replaced the inner cross-validation with a simple split between the training fold and the validation fold (Table S35). Although the hyperparameters were already tuned, a validation set was still required for two reasons: (1) to perform early stopping (Prechelt, 1998), a form of regularization. (2) to generate a set of validation predictions that are necessary for efficient ensemble building (see Methods - Models ensembling) and model selection. During the cross-validation, we scaled and centered the target variable (chronological age) as well as the side predictors (sex and ethnicity) around zero with a standard deviation of one, using the training summary statistics. Scaling the target and the input helps prevent the issues of exploding and vanishing gradients (Hochreiter, 1991; Hochreiter et al., 2001).

##### Cross-validation example

For the sake of clarity, let us walk through an example. Let us say that we want to generate unbiased predictions for every sample in a dataset using a CNN. First, we select the data fold #0 as the validation set, the data fold #1 as the testing set, and the remaining data folds (#2-9) as the training set. Then we scale and center the target (age), and the side predictors (sex and ethnicity) using the training mean and standard deviation: for each of the variables, we subtract the training mean to the variable on both the training, the validation and the testing set, and we divide it by the training standard deviation. We then train the model on the training set until convergence and select the architecture’s parameters (also known as “weights”) associated with the epoch that yielded the lowest validation RMSE. We then use the optimal weights to generate validation predictions for the data fold #0 and testing predictions on the data fold #1. Finally, we unscale the validation and testing predictions by multiplying them by the initial age training standard deviation before adding the initial age training mean to them. This completes the first cross-validation fold.

We then reiterate the process, this time using the data fold #1 as the validation set, the data fold #2 as the testing set, and the remaining data folds (#0 and #3-9) as the training set. We use the optimized weights to generate the validation predictions on the data fold #2, and the testing predictions on the data fold #3. We unscale the validation and testing predictions. This completes the second cross-validation fold. We reiterate the process eight more times to complete the cross-validation. We then concatenate the validation predictions from the ten data folds to obtain the final validation predictions, and the testing predictions from the ten data folds to obtain the final testing predictions.

### Generating average predictions for each participant

We generated an average prediction for each individual, reported to UKB’s instance 0. We walk through an example. Let us assume a participant had two autorefraction samples collected from them in instances 2 and 3, respectively at age 70 and 80. Let us assume that the age predictions were respectively 64 and 78, so the residuals are respectively -6 years and -2 years, for an average of -4 years. However, we still need to take into account the bias in the residuals, defined as the difference between the participant’s chronological age and the prediction. As explained in more details under Methods - Biological age definition, we observed a bias in the residuals as a function of chronological age. Participants on the younger end of the chronological age distribution tend to be predicted older than they actually are, whereas participants on the older end of the distribution tend to be predicted younger than they actually are. We need to properly account for this bias when translating a prediction from a more recent instance to an older instance. Let us assume that the average bias in the residuals for participants who are 70 and 80 years old is respectively -2 years and -4 years. After correcting for this bias, the predictions are now respectively 64-(−2)=66 and 78-(−4) = 82. Therefore, the corrected residuals for this participant are respectively -4 years and +2 years, for an average of -1 years. Finally, let us assume that the participant was 60 years old in instance 0. We will assign a single prediction of 60-1=59 years to the participant, but we still need to un-correct for the bias in residuals. Let us assume that the average bias for the residuals at age 60 is +5 years. We will assign a final prediction for the participant of 59+5=64 years. This new set of predictions reported on the instance 0 is more likely to have a non-zero sample size overlap with other predictors based on datasets collected on instance 0 (e.g. OCT images) and can therefore be leveraged by the ensemble builder.

A key point we would like to highlight here is that we did not actually correct for the bias in the residuals at this step of the pipeline. Instead, we corrected then un-corrected the predictions that we translated from different instances to the instance 0. The actual correction for the residual biases takes place when defining the biological age phenotypes (see Methods - Biological age definition).

To distinguish between raw predictions on the instance 0, and the average predictions reported to the instance 0, we created a new instance which we named instance “*”. We refer to these predictions as “participants predictions”, as opposed to “samples predictions”.

### Genome-wide association study of eye aging

We used the average accelerated aging value over the different samples collected over time (see Supplementary Methods - Generating average predictions for each participant). Next, we performed genome wide association studies [GWASs] in each eye dimension using BOLT-LMM (Loh et al., 2018, 2015b) and estimated the the SNP-based heritability for each of our biological age phenotypes, and we computed the genetic pairwise correlations between dimensions using. We used the v3 imputed genetic data to increase the power of the GWAS, and we corrected all of them for the following covariates: age, sex, ethnicity, the assessment center that the participant attended when their DNA was collected, and the 20 genetic principal components precomputed by the UKB. We used the linkage disequilibrium [LD] scores from the 1,000 Human Genomes Project (Consortium and The 1000 Genomes Project Consortium, 2015). To avoid population stratification, we performed our GWAS on individuals with White ethnicity.

#### Identification of SNPs associated with accelerated eye aging

We identified the SNPs associated with accelerated eye aging dimensions using the BOLT-LMM (Loh et al., 2018, 2015b) software (p-value of 5×10^8^). The sample size for the genotyping of the X chromosome is one thousand samples smaller than for the autosomal chromosomes. We therefore performed two GWASs for fundus and OCT aging, where we (1) excluding the X chromosome, to leverage the full autosomal sample size when identifying the SNPs on the autosome and (2) including the X chromosome, to identify the SNPs on this sex chromosome. We then concatenated the results from the two GWASs to cover the entire genome, at the exception of the Y chromosome.

We used the FUnctional Mapping and Annotation (FUMA) tool for identifying the independent loci of the GWAS results and mapping them to their closest genes (Watanabe et al., 2017) as we recently performed in a prior study on another organ dimension (Le Goallec et al., 2022). Specifically, we identify (1) the loci associated with each of the traits, and the (2) nearest protein coding genes. We have also provided public links to the FUMA analyses, located here: . Briefly, to identify the significant locus, SNPs are filtered that have a GWAS-level of significance (in our study, p < 5e-8). SNPs that are GWA-significant and have a r^2^ greater than 0.6 are candidate SNPs for a locus; other SNPs are considered independent. The SNP with the lowest p-value and independent of other SNPs at a r^2^ less than 0.1 is the “lead SNP”. We report the lead SNP and the number of other SNPs in linkage with the lead SNP. Next, to identify closest protein coding genes, FUMA uses ANNOVAR (Wang et al., 2010), to positionally map SNPs.

We conducted a few steps for GWAS quality control (QC). First, we restricted the GWAS to have INFO scores greater than 0.3 and minor allele frequency of 0.001. The INFO score measures the certainty of the imputation and ranges from 0 (zero certainty) to 1. Imputation was performed by UK Biobank. Second, we estimated the lambda GC in different bins of the INFO and minor allele frequency spectrum.

#### Heritability and genetic correlation

We estimated the heritability of the accelerated aging dimensions using the BOLT-REML (Loh et al., 2015a) software. We included the X chromosome in the analysis and corrected for the same covariates as we did for the GWAS. Using the same software and parameters, we computed the genetic correlations between accelerated aging in the two image-based eye dimensions.

We computed the genetic correlation between age-related macular degeneration [AMD] and accelerated eye aging in each eye dimension in two ways. First, we directly leveraged the AMD cases in the UKB cohort (union of field IDs 6148-5 and ID20002-1528; 5,000 AMD-positive participants out of 502,000) to compute the correlation from participant’s data with BOLT-REML. The limitation of this method is the small sample size of AMD-positive participants in UKB, especially after taking the intersection with participants for which eye images were collected (effective sample size ∼500 for general eye aging). Second, we used LD score regression [LDSC] (B. Bulik-Sullivan et al., 2015; B. K. Bulik-Sullivan et al., 2015) and AMD GWAS summary statistics (Winkler et al., 2020). The limitation of this second approach is that it tends to underestimate heritability since it cannot leverage the full information at the participant level.

### Non-genetic correlates of accelerated eye aging

Each variable was (1) We define as biomarkers the scalar variables measured on the participant, which we initially leveraged to predict age (e.g. blood pressure, Table S10). (2) We define clinical phenotypes as other biological factors not directly measured on the participant, but instead collected by the questionnaire, and which we did not use to predict chronological age. For example, one of the clinical phenotypes categories is eyesight, which contains variables such as “wears glasses or contact lenses”, which is different from the direct refractive error measurements performed on the participants, which are considered “biomarkers” (Table S13). (3) Diseases include the different medical diagnoses categories listed by UKB (Table S16). (4) Family history variables include illnesses of family members (Table S19). (5) Environmental variables include alcohol, diet, electronic devices, medication, sun exposure, early life factors, medication, sun exposure, sleep, smoking, and physical activity variables collected from the questionnaire (Table S22). (6) Socioeconomic variables include education, employment, household, social support and other sociodemographics (Table S25).

### Interpretability of the predictions

#### Scalar data-based predictors

For elastic nets, we interpreted the models using the values of the regression coefficients. Large absolute values for these coefficients means they played an important role when generating the predictions. For gradient boosted machines we used the feature importances, which are based on the number of times a tree selected each of the variables. Variables with high feature importances were selected more often and are therefore likely to play a key role in predicting chronological age. For neural networks, we estimated the importance of each feature by permuting it randomly between samples before computing the performance of the model. The score of each feature is the difference between the R-Squared value before and after the random permutations. Features whose random permutation leads to a large decrease in the model’s performance are estimated to be important predictors of chronological age.

We estimated the concordance between the three different algorithms by computing the Pearson and the Spearman correlations between their feature importances.

#### Image-based predictors

To interpret the CNNs built on images, we first used saliency maps (Alqaraawi et al., 2020), which we coded using the keract python library. For each input sample, a saliency map uses the gradient of the final prediction with respect to each individual input pixel to estimate whether changing the value of this pixel would affect the prediction. Pixels for which the gradient is close to zero are not important, whereas pixels with a large gradient are estimated to be important.

We then built a second attention map using a custom version of the Gradient-weighted Class Activation Mapping [Grad-CAM] algorithm (Selvaraju et al., 2017) adapted to regression rather than multi-class classification: Gradient-weighted Regression Activation Mapping [Grad-RAM]. The intuition behind Grad-CAM maps is that they are similar to saliency maps (Selvaraju et al., 2017), but instead of computing the gradient with respect to the input image, they compute it with respect to the activation of the last convolutional layer. As convolutional layers maintain the spatial organization of the input image, Grad-CAM can still identify which region of the image is driving the predictions. Because Grad-CAM does not have to backpropagate the gradient all the way back to the input image, it is considered a less noisy alternative to the saliency maps. In the same way that saliency maps need to combine the attention maps generated in the different input channels (e.g. RGB) into a single activation map, Grad-CAM must combine the attention maps generated on the different filters of the last convolutional layer. For example, the last convolutional layer for InceptionResNetV2 has 1,792 filters. Grad-CAM combines these 1,792 attention maps into a single attention map using a linear combination. In the initial Class Activation Mapping [CAM] algorithm (Zhou et al., 2016), generating CAM activation maps required to retrain the model after modifying the architecture and replacing all the fully connected layers after the final convolutional layer with a global max pooling operation, which converted each filter into a scalar feature. The intuition behind this substitution was that each filter could be interpreted as detecting a specific feature, and global max pooling yielded a scalar that could be interpreted as the presence (high value) or absence (low value) of the feature anywhere on the image. The scalar values were then linearly combined and activated using the softmax function to yield the probabilities of belonging to different classes. To obtain the activation map for a specific class, the filters of the last convolution layer were linearly combined using the weights connecting the scalar features obtained after the max pooling operation to the final prediction score for that class. CAM was later improved to become Grad-CAM (Selvaraju et al., 2017). Grad-CAM saves the need for modifying the architecture of the model and retraining it by approximating the linear regression weight for each final convolutional filter by the mean activation gradient over the pixels of the filter. The intuition behind this approximation is that a filter’s pixel is important if changing its value affects the final prediction, so a high average gradient over the pixels of the filter justifies that this filter should be given a higher weight when merging all the filters into a single attention map. To adapt Grad-CAM to our regression task we (1) computed the derivatives of the chronological age prediction rather than a class’ prediction and (2) removed the ReLU activation applied to the weighted sum of the last convolutional filters, which we replaced by an absolute value. The rationale is that for (Grad-)CAM maps, we only want to highlight the regions of the picture which are associated with a high probability for the class. In contrast, for (Grad-)RAM we care as much about the regions of the input image that can strongly increase the chronological age prediction as about the regions that can strongly decrease it. Because the filters in the last convolutional layer are the result of the processing of the input image by several convolutional layers with possibly negative weights, the sign of the last convolutional layer’s pixels and regression weights cannot be linked to either accelerated aging or decelerated aging, only to the magnitude of the shift that would affect the prediction if each region of the input image was modified. Regression Activation Mapping (RAM) was mentioned as a possible extension of CAM in the original CAM publication (Zhou et al., 2016) and has been used to interpret models CNNs built on retinal images (Wang and Yang, 2017) and cortical surfaces (Duffy et al., 2019), but we are to our knowledge the first to describe the generalization of Grad-CAM to a regression task. One notable difference between our implementation and Wang and Yang.’s implementation (Wang and Yang, 2017) is that we are taking the absolute value of the final attention map, as mentioned above. We found that not taking the absolute value led to misleading attention maps for participants with high chronological age predictions. The attention map highlights important areas with negative values, which are therefore depicted in blue, a color otherwise associated with unimportant regions in traditional CAMs. Inversely, regions on the input image for which the attention map has a slight positive value are spuriously considered to be the most important and are highlighted in red. We therefore advise that RAM or Grad-RAM be implemented using an absolute value. We coded Grad-RAM using the get_activations and get_gradients_of_activations functions of the keract python library.

It is important to understand that unlike the feature importances described under “Scalar data-based predictors”, which describe the model itself, attention maps are sample specific. In other words, they can be used to explain which features drove the predictions for a specific inputted sample but cannot provide an explanation for the way the model is performing predictions in general.

For each aging subdimension, we generated the attention maps for the best performing CNN architecture. We selected representative samples for which we computed the different attention maps. We computed attention maps for the two sexes (female and male), for three age ranges (ten youngest ages, ten middle ages and ten oldest ages of the chronological age distribution) and for three aging rates (accelerated agers, normal agers, decelerated agers). For each intersection of the three categories listed above, we selected the ten most representative samples (e.g. the ten most accelerated agers among young males). The figures in this paper only present the first, most representative of these ten samples. The complete set of samples can be found on the website.

### Non-genetic correlates of accelerated aging

Unlike DNA, biomarkers, phenotypes, diseases, family history, environmental variables and socioeconomics can change over life. As a consequence, we compared each biomarker, phenotype and environmental variable with the accelerated aging of the participant at the time the exposure was measured and we used the “Samples predictions”, as opposed to the “Participants predictions” that we used for the identification of genetic correlates (see Methods - Models ensembling - Generating average predictions for each participant).

#### Imputation of the non-genetic X-variables

Most X-variables were not collected on all four instances. Additionally, no X-variables were collected at the same time as the accelerometer data was collected. To identify the non-genetic correlates of accelerated aging, we had to impute the values of the X-variables for the ages of the participants for which they were not available. We considered two imputation methods, which we refer to as the “cross-sectional” and the “longitudinal” imputations.

For the cross-sectional imputation, we computed a linear regression for each X variable as a function of age, adjusting for sex. We then used the slope of the linear regression to extrapolate the value of the XWAS variable at different ages.

For the longitudinal imputation, we first selected, for each X variable, all the participants that had at least two measures taken for this X variable. We then performed a linear regression for each participant. We then averaged the slope of the linear regressions over all the participants of the same sex. Finally, we used this slope to extrapolate the value of the XWAS variable at different ages for all participants depending on their sex, in the same way we did it for the cross-sectional imputation.

It is important to notice that for both the cross-sectional imputation and the longitudinal imputation, data can only be imputed when the XWAS variable has been measured at least once for the participant. This raw measure is then used to extrapolate which value the X variable was likely taking a couple years earlier and/or later.

The advantage of the cross-sectional imputation is larger sample sizes. The advantage of the longitudinal method is that it corrects for generational effects. For example, old people have shorter legs than young people on average (Le Goallec and Patel, 2019). This is not because human legs shrink as we grow older. Instead, people who are old today already had shorter legs when they were young. If the cross-sectional regression is used to impute the length of the participants on instances where it was not measured, it will spuriously assign smaller values to the older samples. In contrast, the longitudinal regression learns the regression coefficient by comparing each participant to themselves as they age and will therefore not capture the generational effect. When used to predict the participants legs’ length, it will impute constant values over time. To evaluate which of the two imputation methods should be preferred, we used them to predict X-variables for which we knew the actual values and computed the R-Squared values associated with the predictions. We found that, even with sample sizes as small as 200 samples, longitudinal imputation outperformed cross-sectional imputation. We therefore used longitudinal imputation.

#### X-Wide Association Studies

First, we tested for associations in an univariate context by computing the partial correlation between each X-variable and eye aging dimensions. To compute the partial correlation between an X-variable and an aging, we followed a three steps process. (1) We ran a linear regression on each of the two variables, using age, sex and ethnicity as predictors. (2) We computed the residuals for the two variables. (3) We computed the correlation between the two residuals and the associated p-value if their intersection had a sample size of at least ten samples. We used a threshold for significance of 0.05 and corrected the p-values for multiple testing using the Bonferroni correction. We plotted the results using a volcano plot. We refer to this pipeline as an X-Wide Association study [XWAS].

In the supplementary tables and the results, we rank the X-variables subcategories by decreasing percentage of variables associated with accelerated aging (note that the ranking is therefore biased towards categories with fewer variables). For each subcategory, we list the three most associated variables, based on the absolute value of the correlation coefficient. For the exhaustive list, please refer to https://www.multidimensionality-of-aging.net/xwas/univariate_associations.

#### Prediction of accelerated eye aging

We leveraged the pipeline we built to predict chronological age as a function of scalar biomarkers to predict accelerated aging for the different eye aging dimensions as a function of the biomarkers, clinical phenotypes, diseases, family history, environmental and socioeconomic variables. We leveraged the same pipeline to identify which features were driving the predictions. We built a model for each X-variables subcategory (Table S10, Table S13, Table S16, Table S19, Table S22, Table S25).

#### X-Correlations between the eye aging dimensions

##### X-Correlations based on the XWAS results

For the sake of clarity, let us walk through an example. We want to compute the environmental correlation between accelerated fundus-based and OCT-based eye aging. The XWAS generates a vector whose components are the partial correlations between the accelerated aging phenotype and each environmental variable, for both fundus-based and OCT-based eye aging. We compute three different Pearson correlations between these two partial correlation vectors. (1) The “All” correlation, using all the components of the two vectors; this correlation tends to be inflated by the large number of X-variables whose correlation with both accelerated aging dimensions is close to zero. (2) The “Intersection” correlation, using only the environmental variables that were significantly associated with both of the accelerated aging dimensions;because the cardinality of the intersection can be small, a small number of X-variables can yield very high or very low correlations. (3) The “Union” correlation, using only the environmental variables that were significantly associated with at least one of the two accelerated aging dimensions; the “Union” correlation represents a compromise between the “All” and the “Intersection” correlations. The figures in this paper were generated using the “Union” correlation, but all three correlations can be explored on the website.

##### X-Correlations based on the feature importances

We then computed the correlations between the feature importances for different accelerated aging dimensions to estimate the X-correlation between the different dimensions. We used the same method as described above under “X-Correlations based on the XWAS results”, replacing the coefficient obtained for each X-variable in a univariate context (using partial correlation with accelerated aging) with the coefficient obtained in a multivariate context (as an accelerated aging predictor in a multivariate model).

## Supplementary Figures

**Figure S1:**
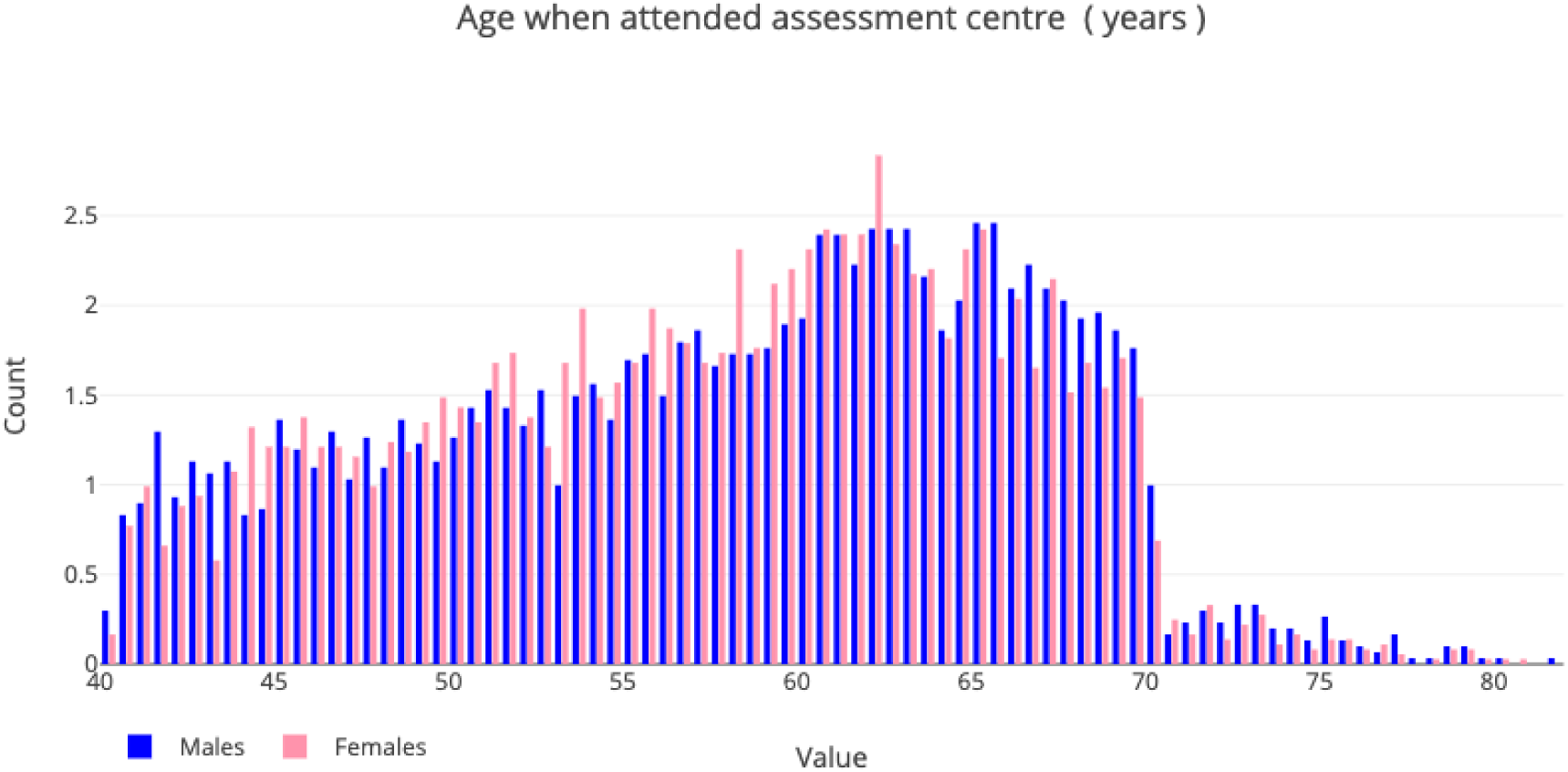
Demographics of the UK Biobank cohort.

**Figure S2:**
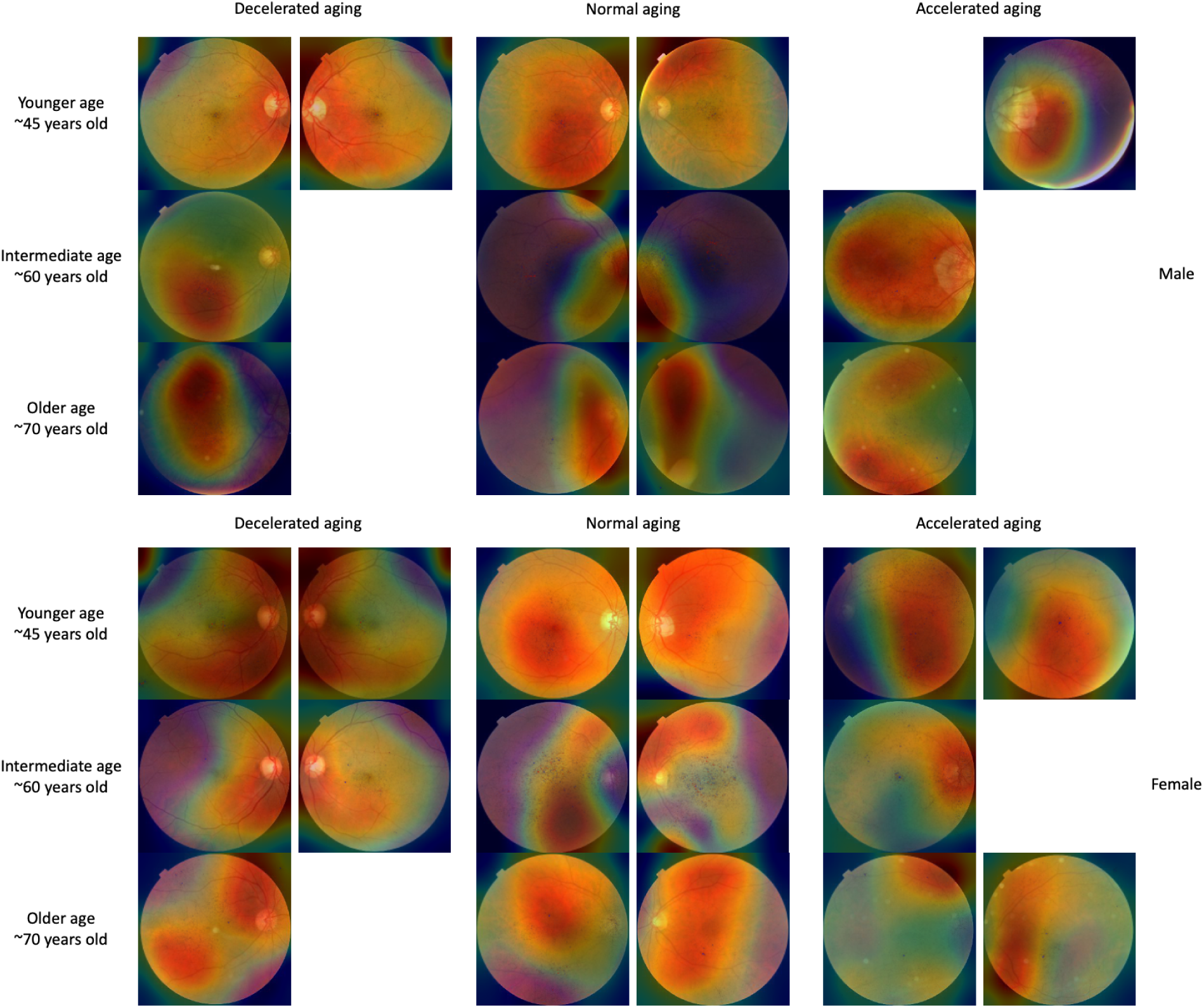
Attention map samples for fundus image-based models. Warm filter colors highlight regions of high importance according to the Grad-RAM map. Missing images are left as a white space.

**Figure S3:**
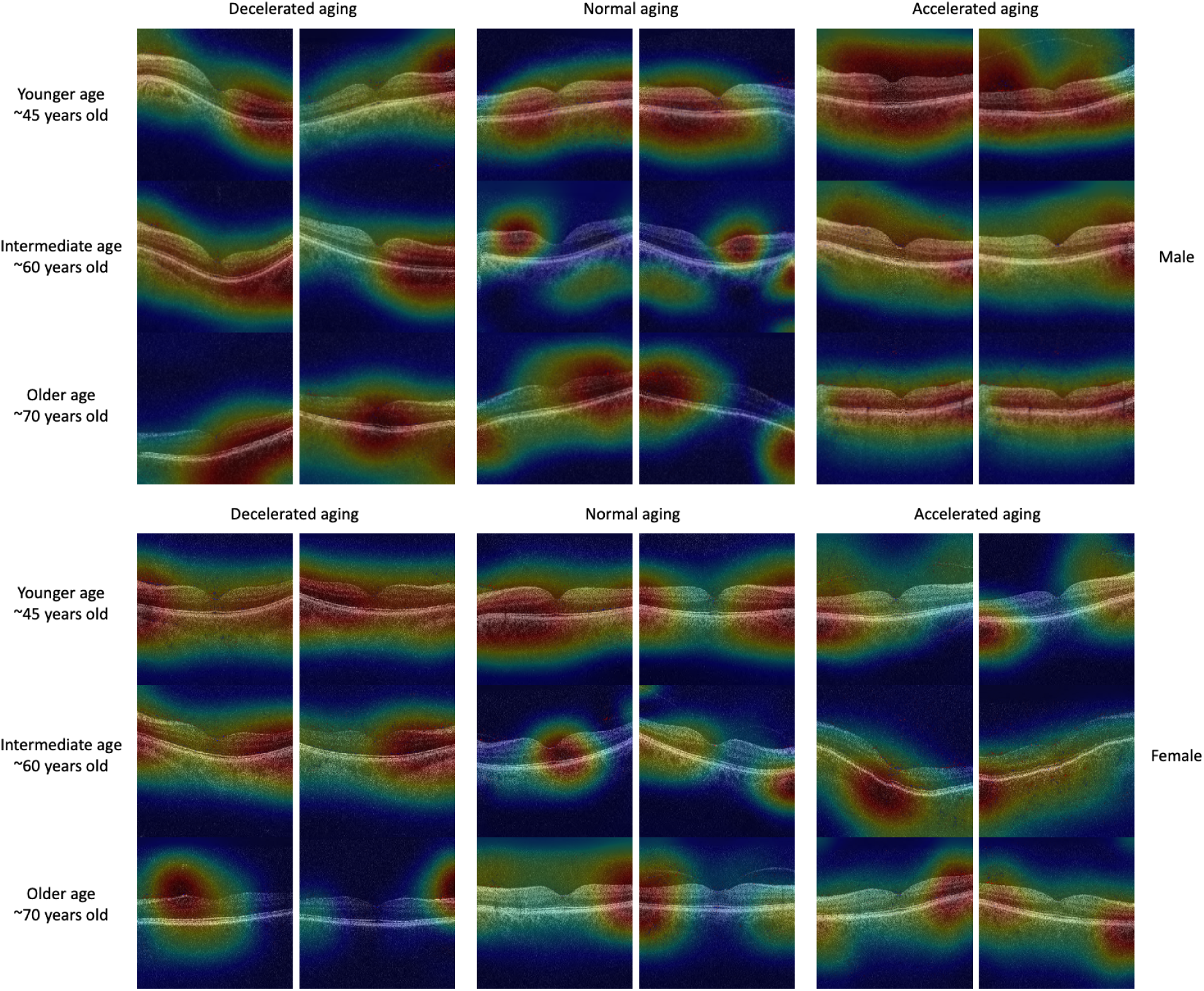
Attention map samples for OCT image-based models. Warm colors highlight regions of high importance according to the Grad-RAM map.

**Figure S4:**
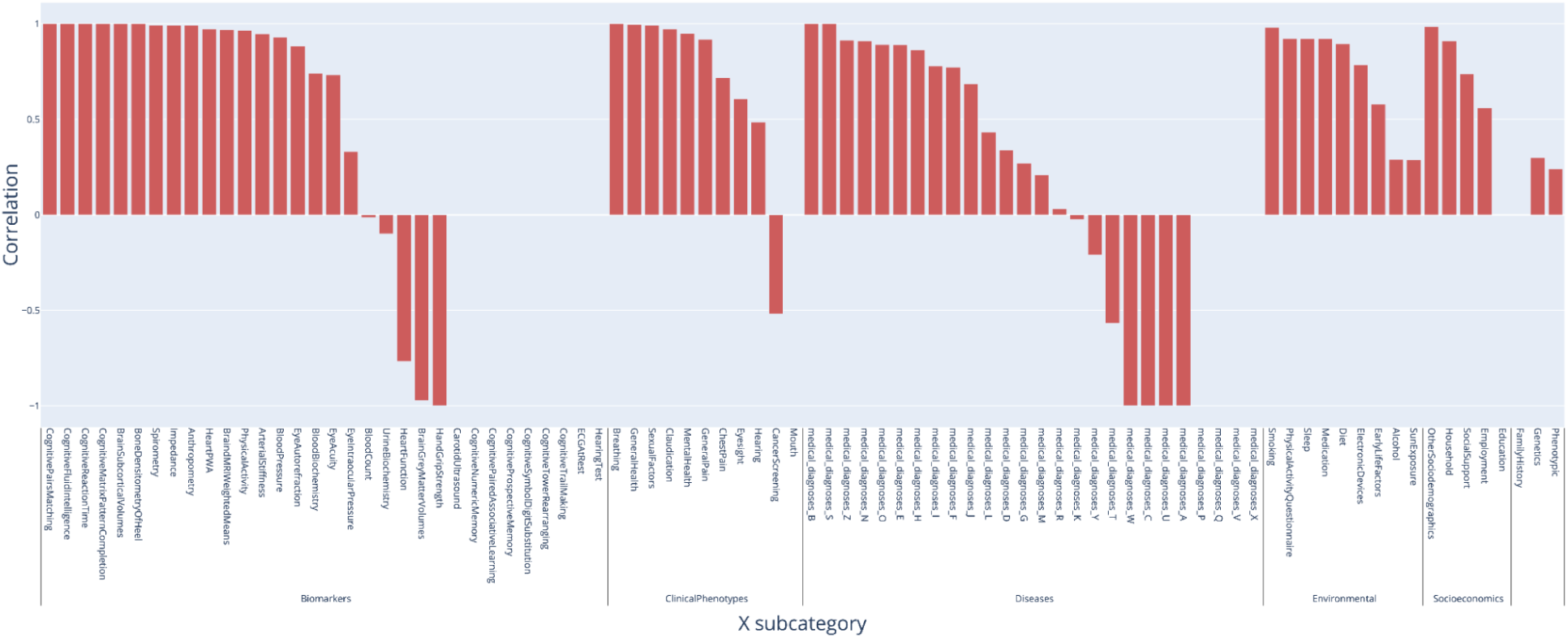
Correlation between fundus-based and OCT-based accelerated eye aging in terms of associated biomarkers, clinical phenotypes, diseases, family history, environmental and socioeconomic variables.

**Figure S5:**
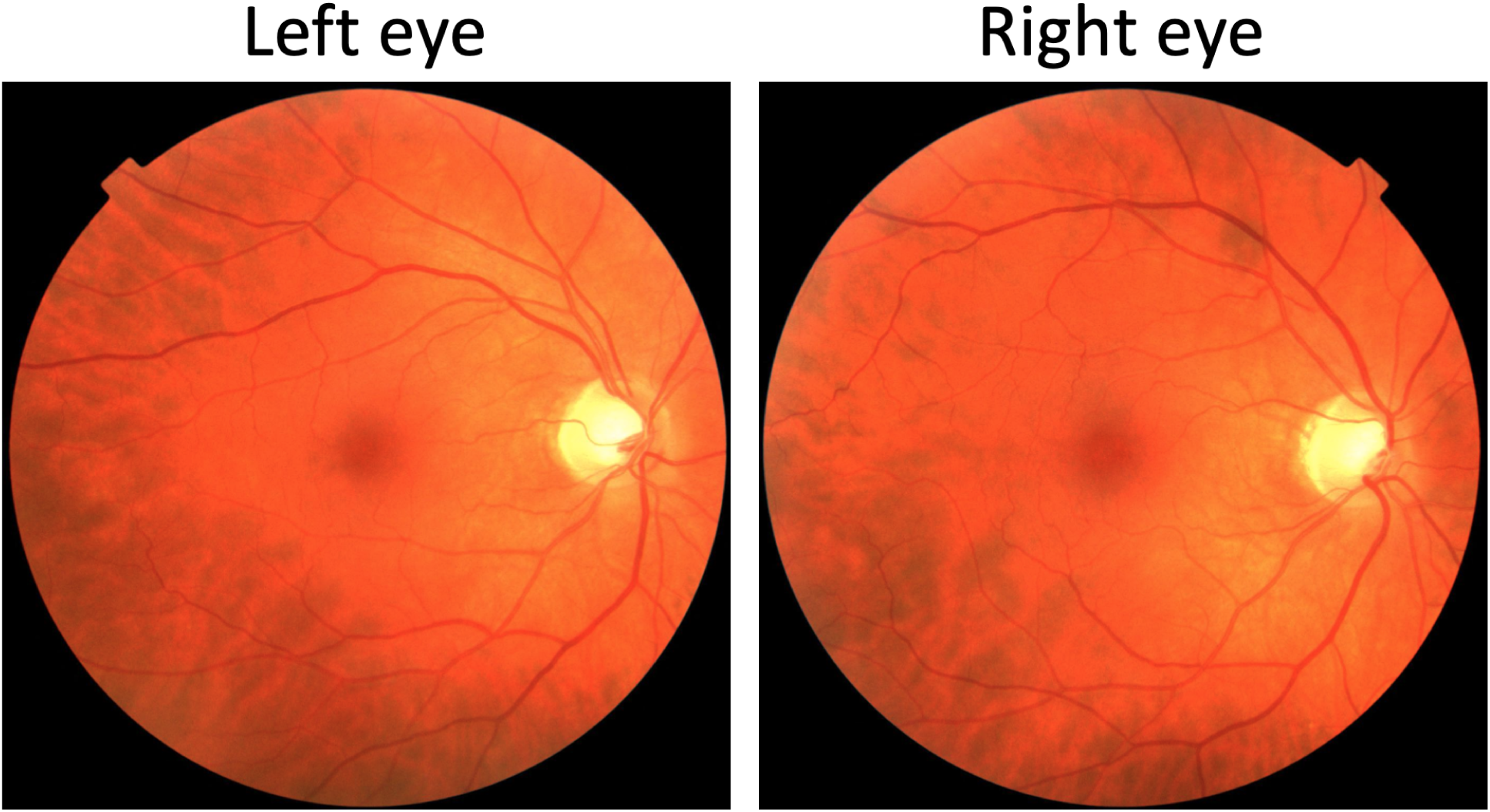
Sample preprocessed left and right eye fundus images. The participant is a 45-50-year-old male.

**Figure S6:**
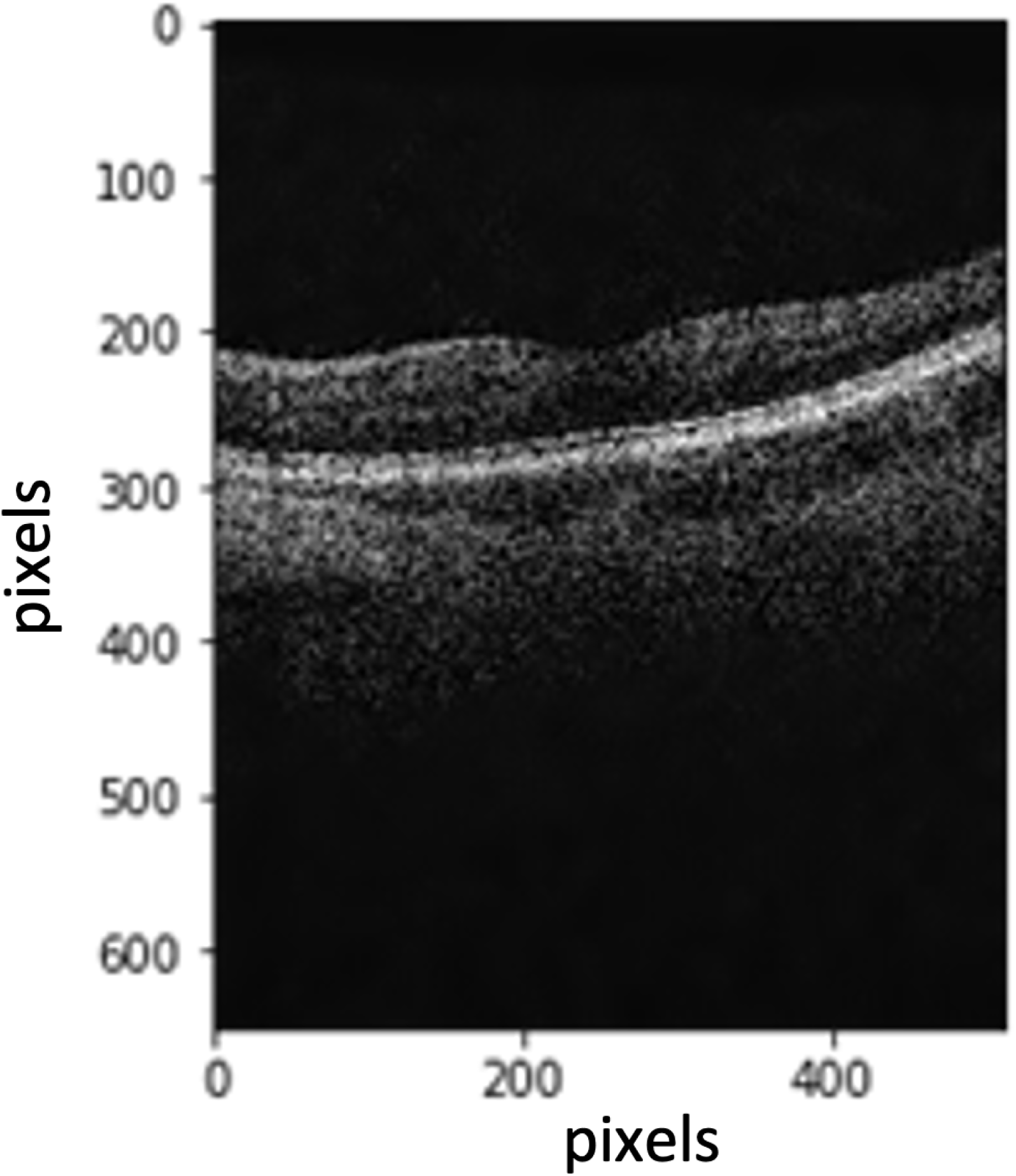
Preprocessing of eye OCT images: raw image.

**Figure S7:**
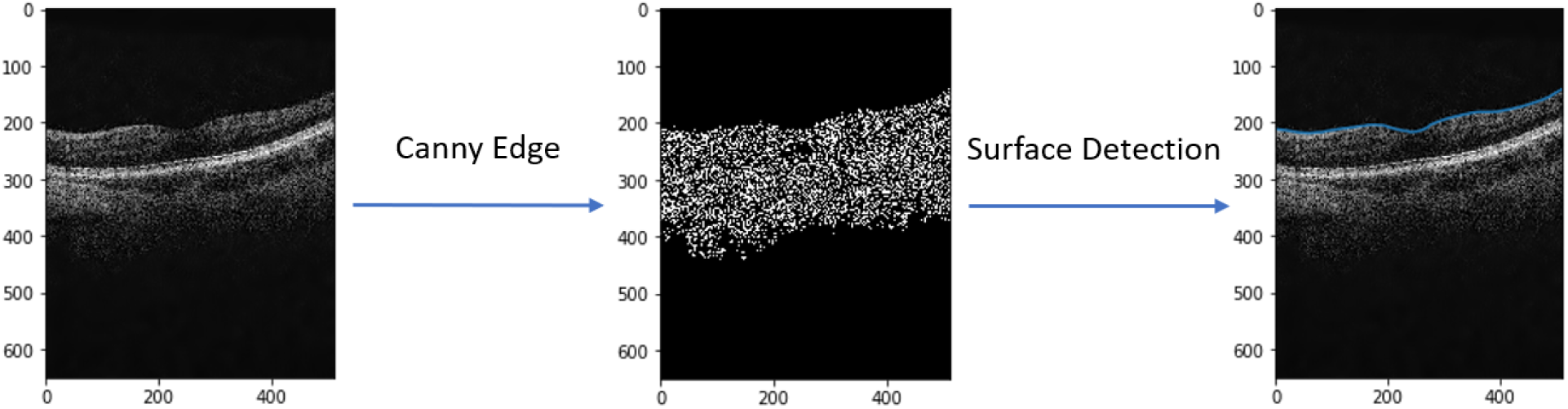
Preprocessing of eye OCT images: detection of the upper surface.

**Figure S8:**
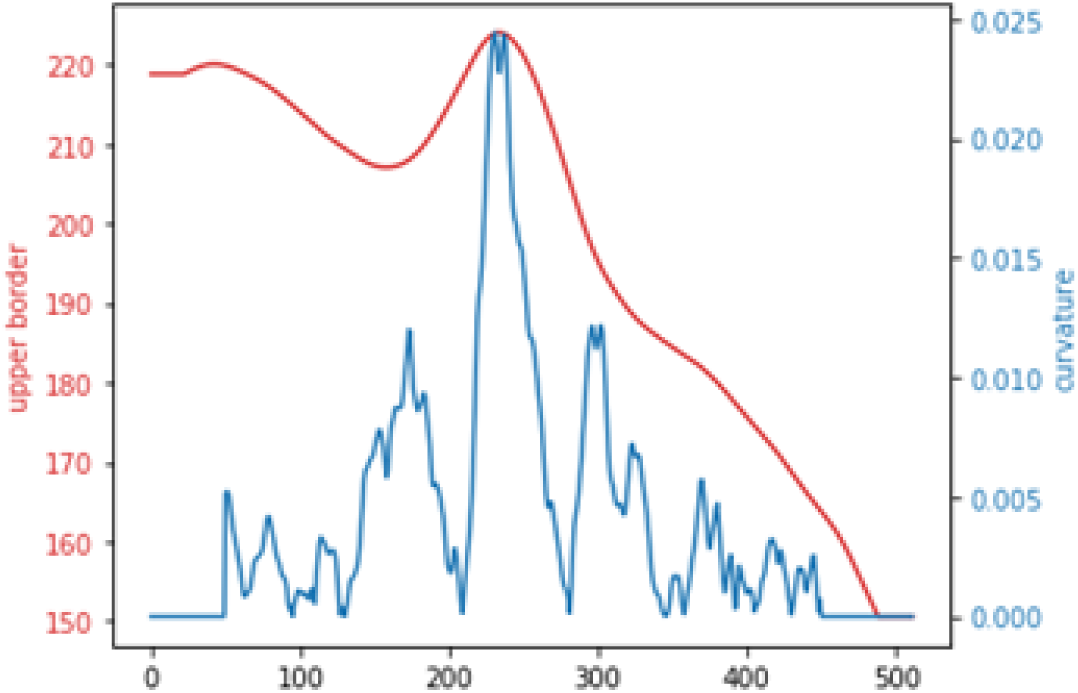
Preprocessing of eye OCT images: computation of the curvature of the upper surface.

**Figure S9:**
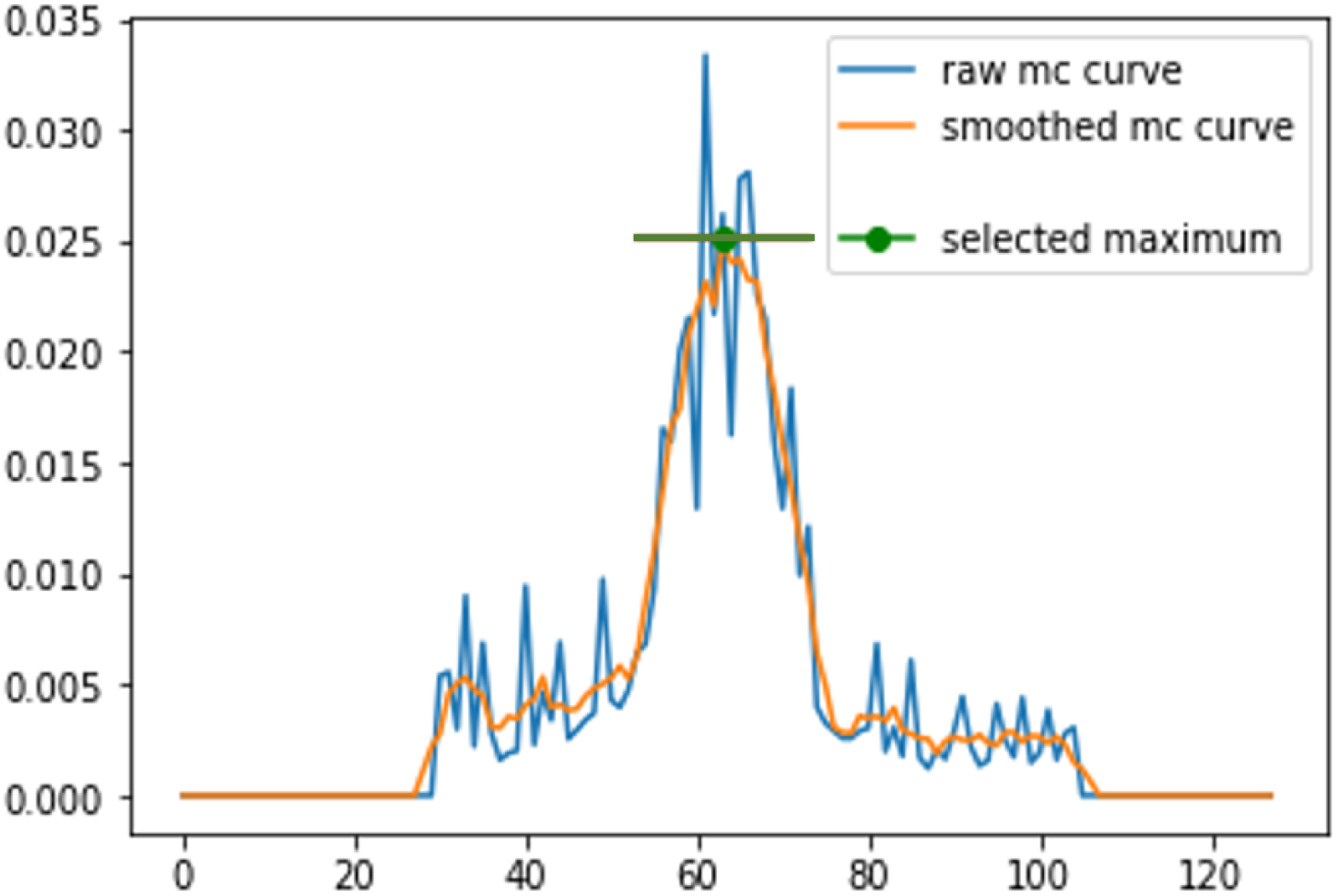
Preprocessing of eye OCT images: detection of image with the maximal curvature after applying the Savitzky–Golay filter. mc stands for maximal curvature.

**Figure S10:**
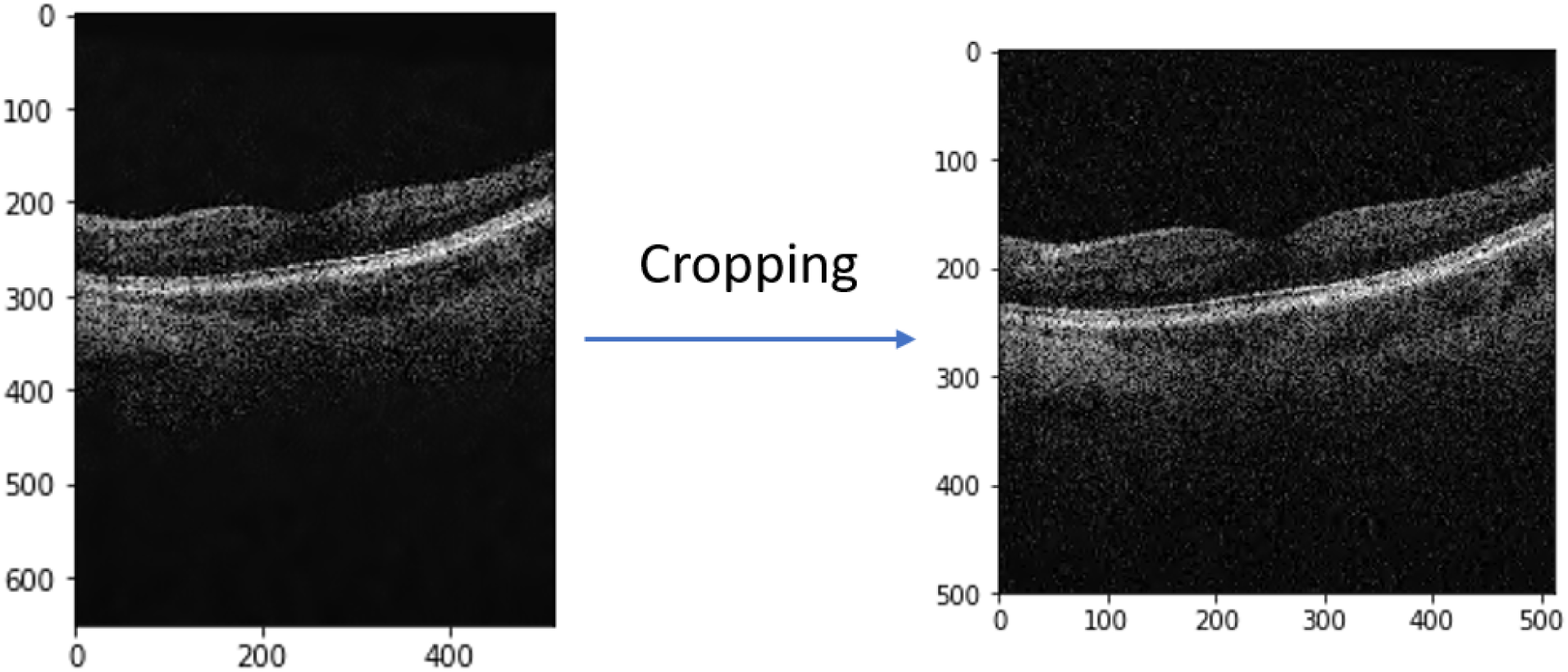
Preprocessing of eye OCT images: raw image vs. cropped image.

**Figure S11:**
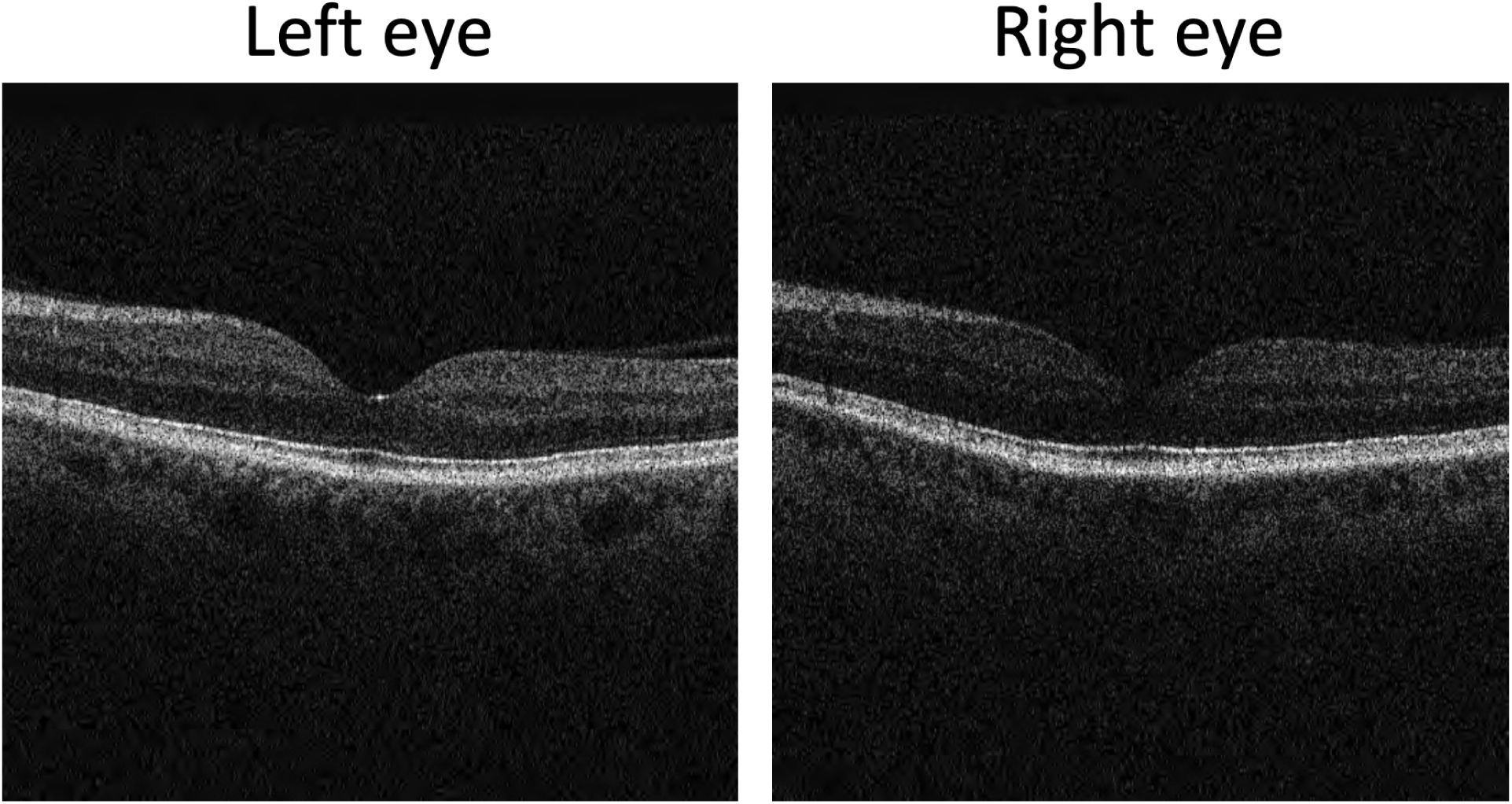
Sample preprocessed OCT left and right eyes images. The participant is a 60-65-year-old male.

**Figure S12.**
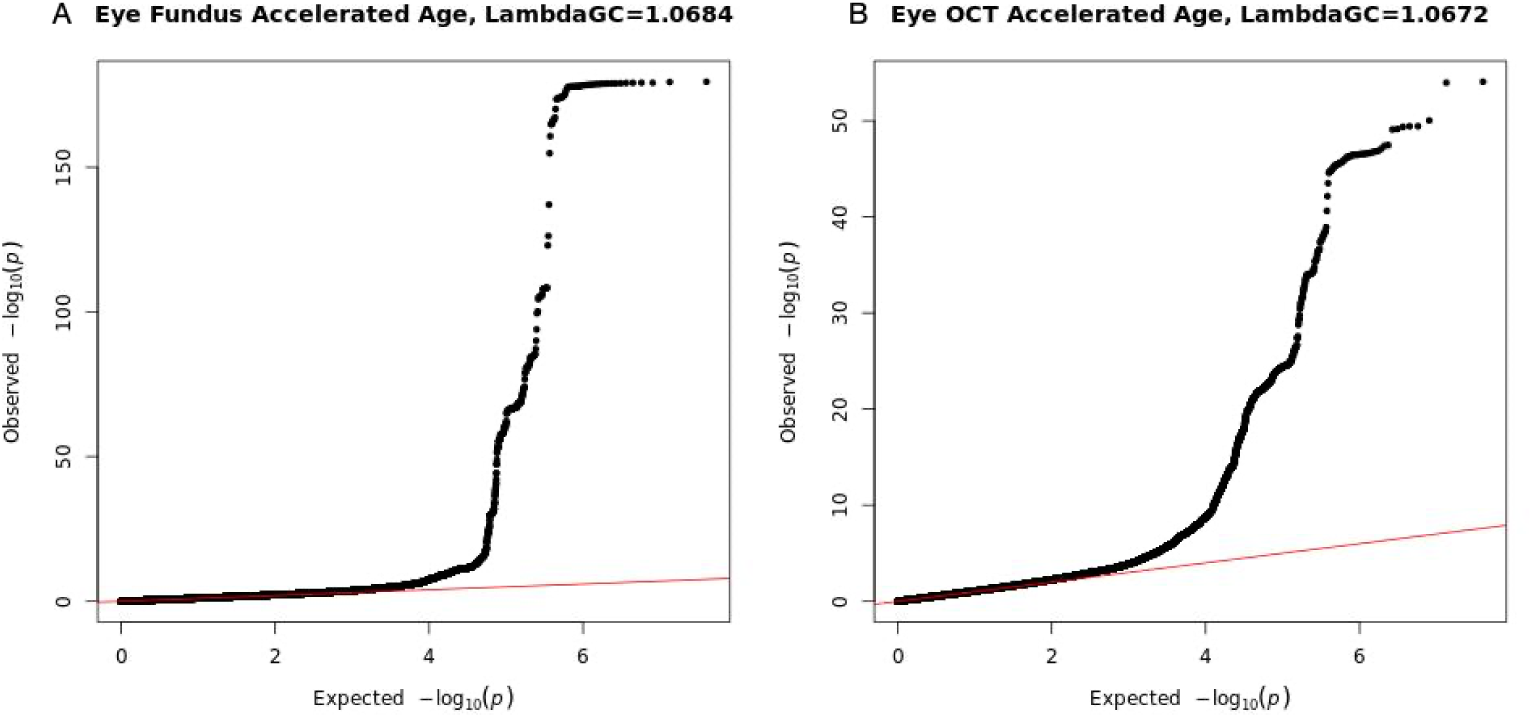
Observed -log10(pvalue) vs. expected -log10(pvalue) for A. fundus-based aging and B. OCT-based aging.

**Figure S13.**
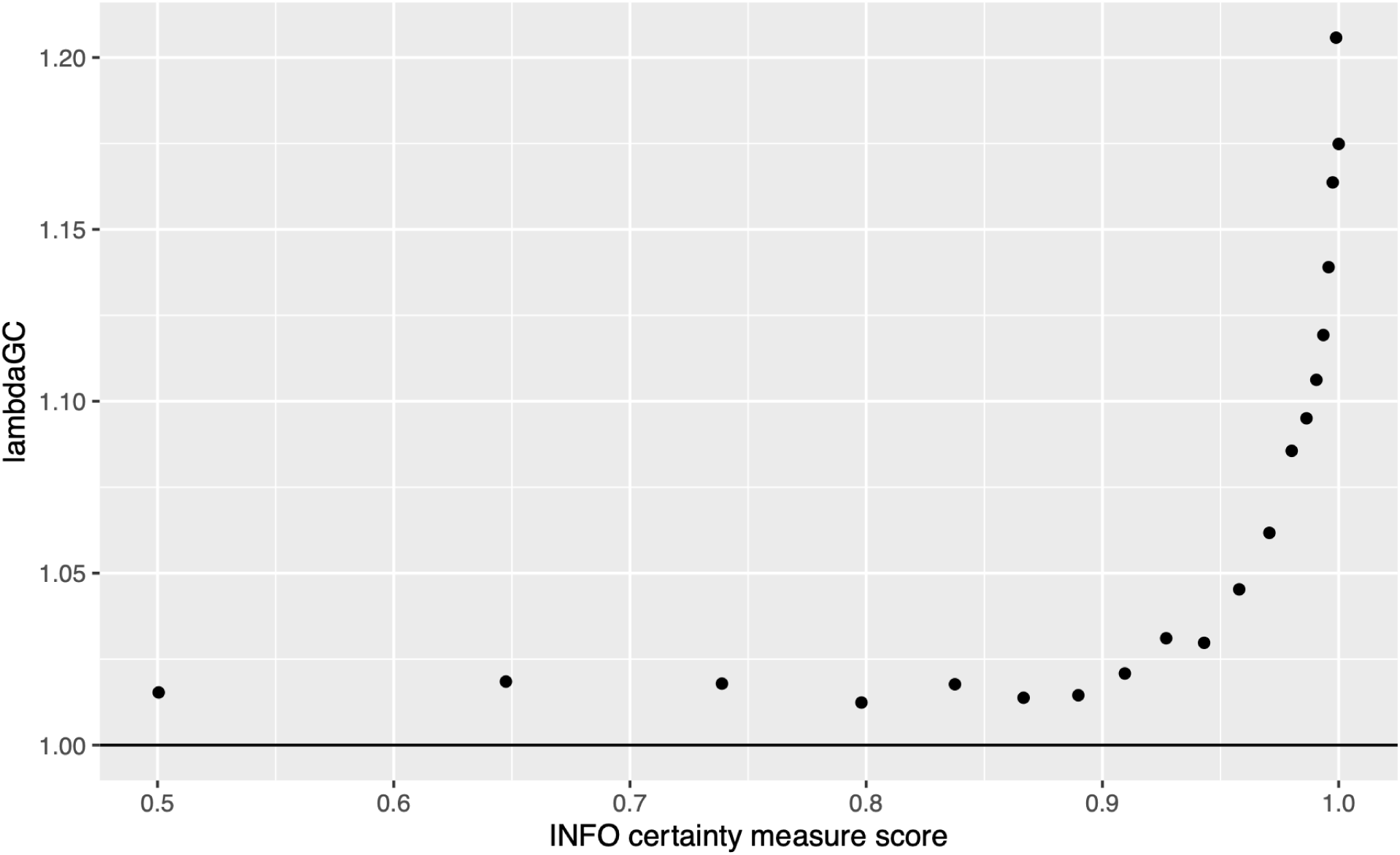
Lambda genetic control versus INFO score. We did not observe higher lambda at poorer imputation quality (or lower INFO score).

**Figure S14.**
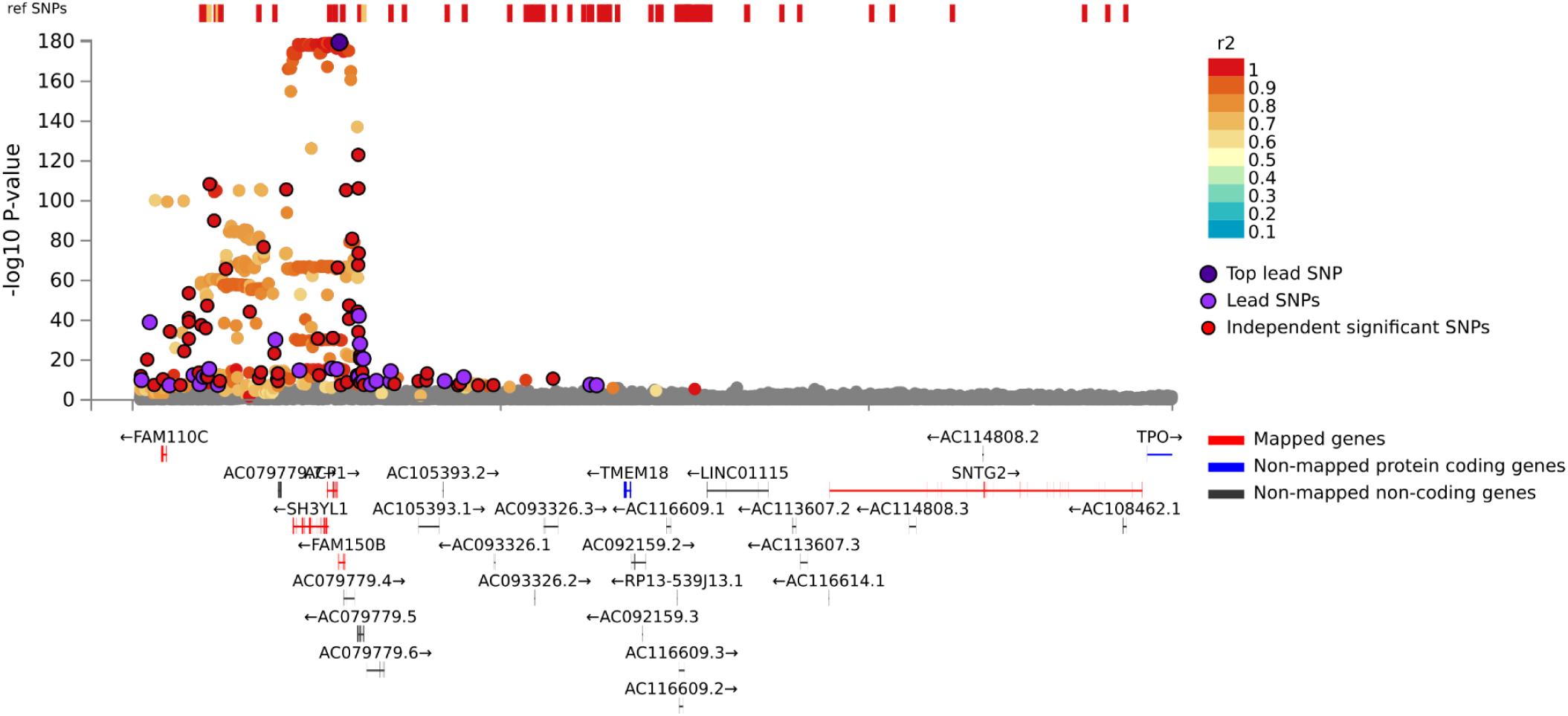
Locus zoom plot of variants around the rs7605824 locus. *FAM150B* is the closest gene.

**Figure S15.**
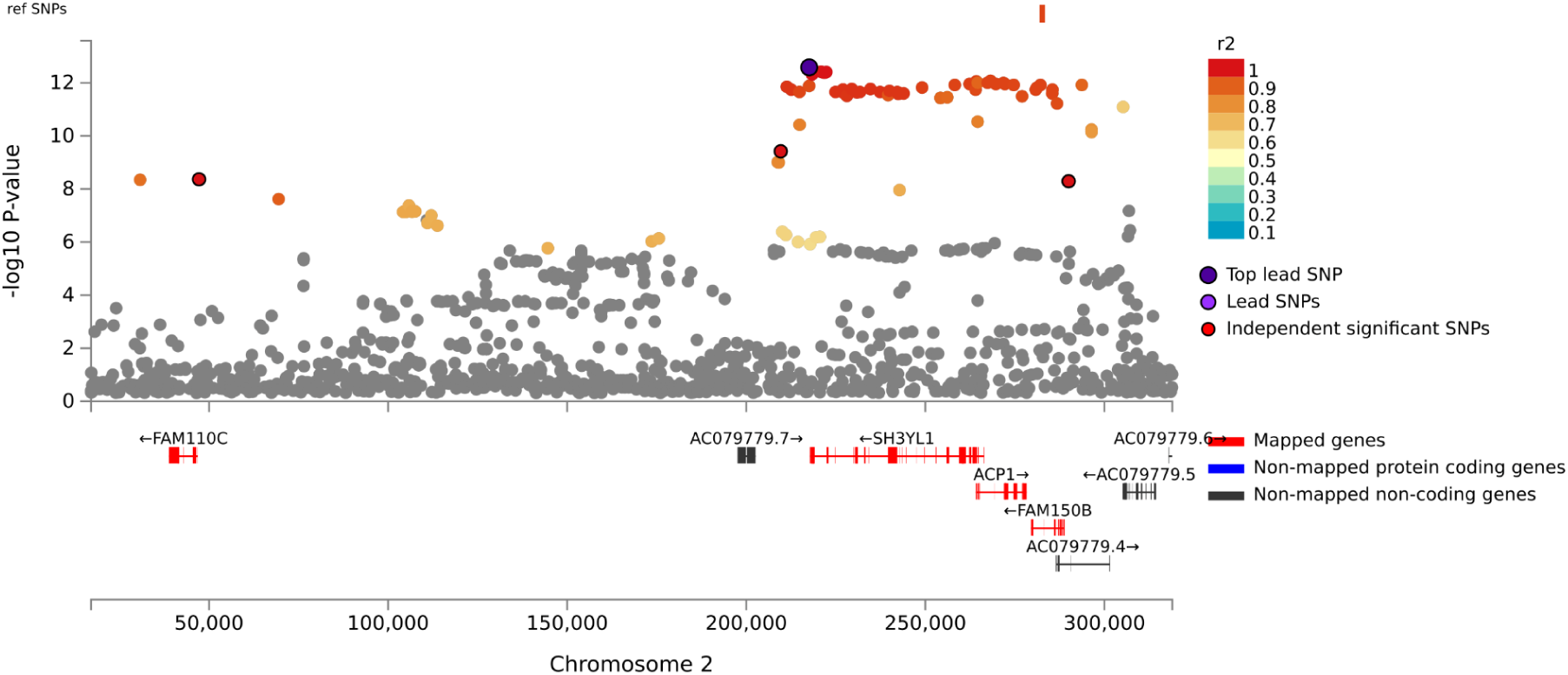
Locus zoom plot of variants around the rs55742348 locus. *SH3YL1* is the closest gene.

## Supplementary Tables

For “See supplementary data”, please refer to: https://www.dropbox.com/s/px2re5qw9n11htt/Supplementary%20data.zip?dl=0

**Table S1:**
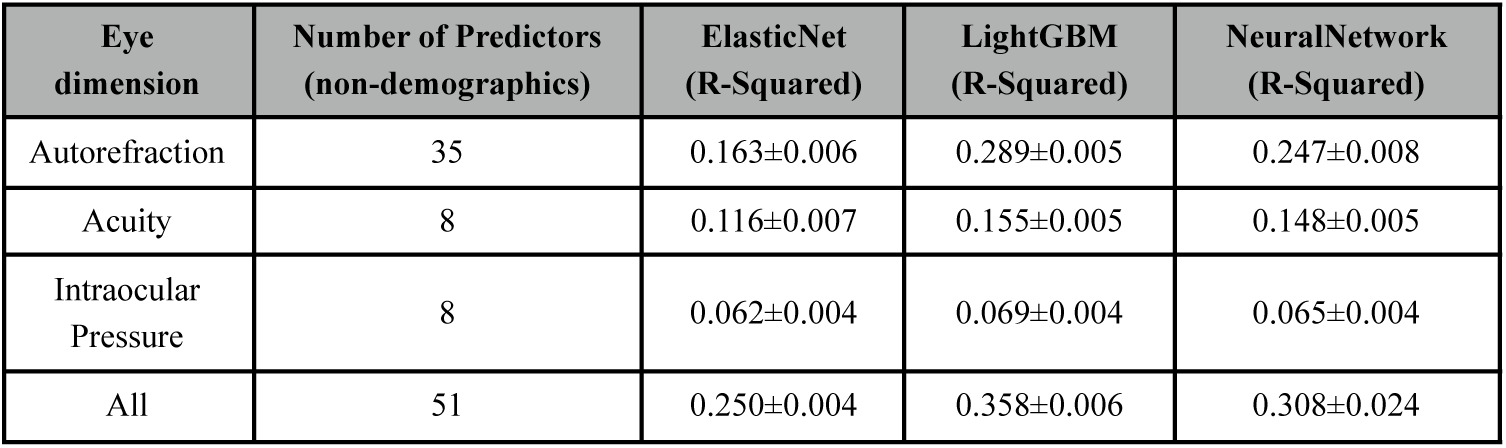
Comparison between the models trained on scalar features.

| Eye dimension | Number of Predictors (non-demographics) | ElasticNet (R-Squared) | LightGBM (R-Squared) | NeuralNetwork (R-Squared) |
| --- | --- | --- | --- | --- |
| Autorefraction | 35 | 0.163±0.006 | 0.289±0.005 | 0.247±0.008 |
| Acuity | 8 | 0.116±0.007 | 0.155±0.005 | 0.148±0.005 |
| Intraocular Pressure | 8 | 0.062±0.004 | 0.069±0.004 | 0.065±0.004 |
| All | 51 | 0.250±0.004 | 0.358±0.006 | 0.308±0.024 |

Table S2: **Feature importances for the models built on all scalar features**

See supplementary data: https://www.dropbox.com/s/px2re5qw9n11htt/Supplementary%20data.zip?dl=0

Table S3: **Feature importances for the models built on autorefraction scalar features**

See supplementary data

Table S4: **Feature importances for the models built on acuity scalar features**

See supplementary data

Table S5: **Feature importances for the models built on intraocular scalar features**

See supplementary data

**Table S6:** Pearson correlations between the feature importances for different scalar features-based algorithms.

| Eye subdimension | Correlation Versus ElasticNet | Correlation Versus LightGBM | Correlation Versus NeuralNetwork | ElasticNet Versus LightGBM | ElasticNet Versus NeuralNetwork | LightGBM Versus NeuralNetwork |
| --- | --- | --- | --- | --- | --- | --- |
| Autorefraction | 0.71 | 0.23 | 0.198 | 0.453 | 0.146 | 0.188 |
| Acuity | 0.803 | 0.61 | 0.541 | 0.878 | 0.394 | 0.147 |
| Intraocular Pressure | 0.4 | 0.296 | 0.633 | 0.521 | 0.888 | 0.389 |
| All Scalars | 0.647 | 0.226 | 0.496 | 0.434 | 0.408 | 0.124 |

**Table S7:** Spearman correlations between the feature importances for different scalar features-based algorithms.

| Eye subdimension | Correlation Versus ElasticNet | Correlation Versus LightGBM | Correlation Versus NeuralNetwork | ElasticNet Versus LightGBM | ElasticNet Versus NeuralNetwork | LightGBM Versus NeuralNetwork |
| --- | --- | --- | --- | --- | --- | --- |
| Autorefraction | 0.745 | 0.154 | 0.323 | 0.397 | 0.304 | 0.295 |
| Acuity | 0.663 | 0.715 | 0.663 | 0.704 | 0.476 | 0.741 |
| Intraocular Pressure | 0.228 | 0.503 | 0.751 | 0.524 | 0.442 | 0.67 |
| All Scalars | 0.641 | 0.226 | 0.488 | 0.317 | 0.424 | 0.302 |

Table S8: **All Single Nucleotide Polymorphisms identified at GWAS-level of significance** rsID, chromosome, position (pos), minor allele frequency (MAF), beta coefficient, standard error, r2 (Linkage Disequilibrium) with independent significant SNP, nearest gene, function of SNP.

**Table S9:** Genetic correlation between accelerated eye aging and age-related macular degeneration.

| Eye dimension | BOLT-REML | LD Score Regression |
| --- | --- | --- |
| General | .614±.421 | .091±.076 |
| Fundus | .172±.142 | .002±.071 |
| OCT | .263±.202 | .138±.129 |

Table S10: **List of biomarkers by subcategories for the Biomarkers Wide Association Study [BWAS]**

See supplementary data

Table S11: **Biomarkers most associated with accelerated aging for each eye aging dimension**

See supplementary data

Table S12: **Biomarkers most associated with decelerated aging for each eye aging dimension**

See supplementary data

Table S13: **List of clinical phenotypes by subcategories for the Clinical Phenotypes Wide Association Study [CWAS]**

See supplementary data

Table S14: **Clinical phenotypes most associated with accelerated aging for each eye aging dimension**

See supplementary data

Table S15: **Clinical phenotypes most associated with decelerated aging for each eye aging dimension**

See supplementary data

Table S16: **List of diseases by subcategories for the Diseases Wide Association Study [DWAS]**

See supplementary data

Table S17: **Diseases most associated with accelerated aging for each eye aging dimension**

See supplementary data

Table S18: **Diseases most associated with decelerated aging for each eye aging dimension**

See supplementary data

Table S19: **List of family history variables by subcategories for the Family History Phenotypes Wide Association Study [FWAS]**

See supplementary data

Table S20: **Family history variables most associated with accelerated aging for each eye dimension**

See supplementary data

Table S21: **Family history variables most associated with decelerated aging for each eye aging dimension**

See supplementary data

Table S22: **List of environmental variables by subcategories for the Environmental Wide Association Study [EWAS]**

See supplementary data

Table S23: **Environmental variables most associated with accelerated aging for each eye aging dimension**

See supplementary data

Table S24: **Environmental variables most associated with decelerated aging for each eye aging dimension**

See supplementary data

Table S25: **List of socioeconomic variables by subcategories for the Socioeconomics Wide Association Study [SWAS]**

See supplementary data

Table S26: **Socioeconomic variables most associated with accelerated aging for each eye aging dimension**

See supplementary data

Table S27: **Socioeconomic variables most associated with decelerated aging for each eye aging dimension**

See supplementary data

Table S28: **Exhaustive XWAS results - association between non-genetic factors and accelerated aging in each eye dimension**

See supplementary data

**Table S29:**
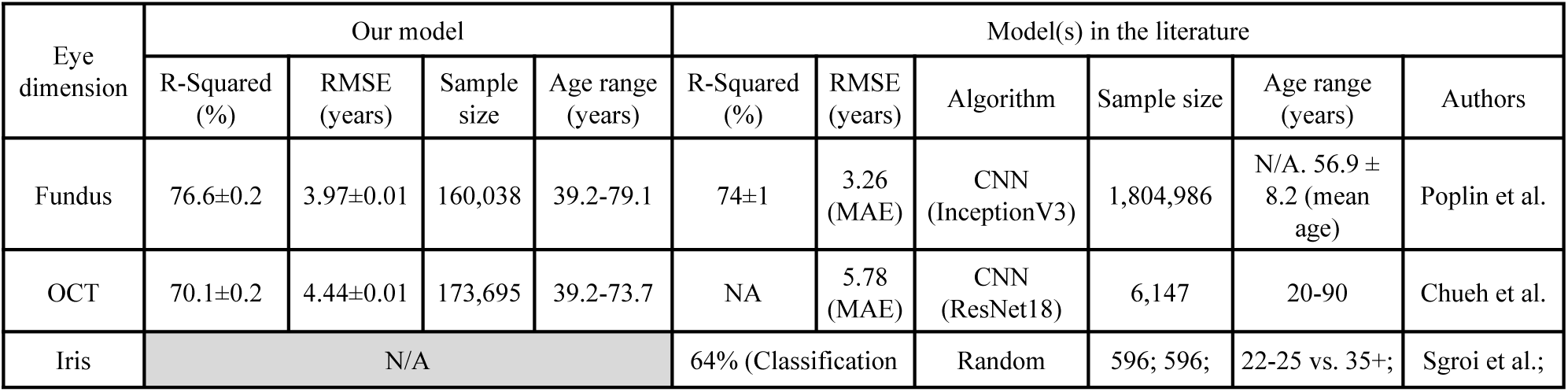
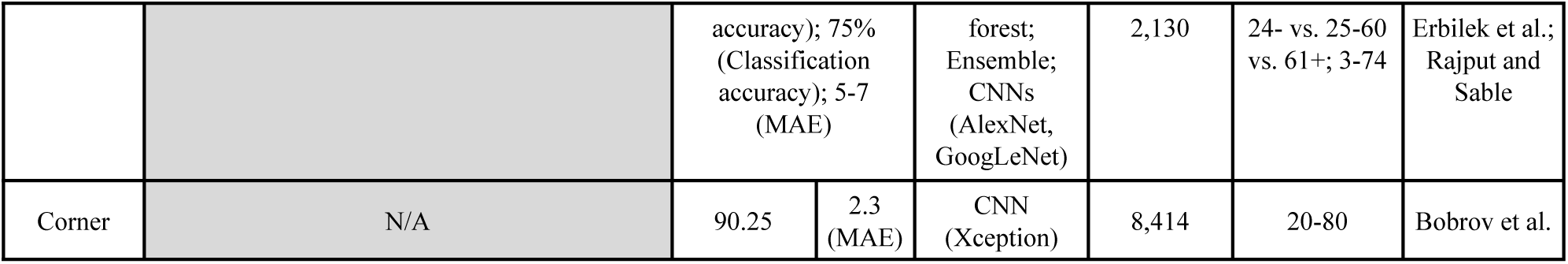
Comparison between our eye age predictors and the literature in terms of prediction performance.

**Table S30:** Image sizes after resizing.

| Eye dimension | Size before<br>resizing | Size after<br>resizing |
| --- | --- | --- |
| Fundus | 1388, 1388 | 316, 316 |
| OCT | 500, 512 | 312, 320 |

**Table S31:** Data augmentation hyperparameters for eye images.

| Dataset | Rotation<br>range | Horizontal shift percentage<br>range | Vertical shift percentage<br>range | Zoom<br>range |
| --- | --- | --- | --- | --- |
| Fundus | 20 | 0.02 | 0.02 | 0 |
| OCT | 30 | 0.1 | 0.2 | 0 |

**Table S32:**
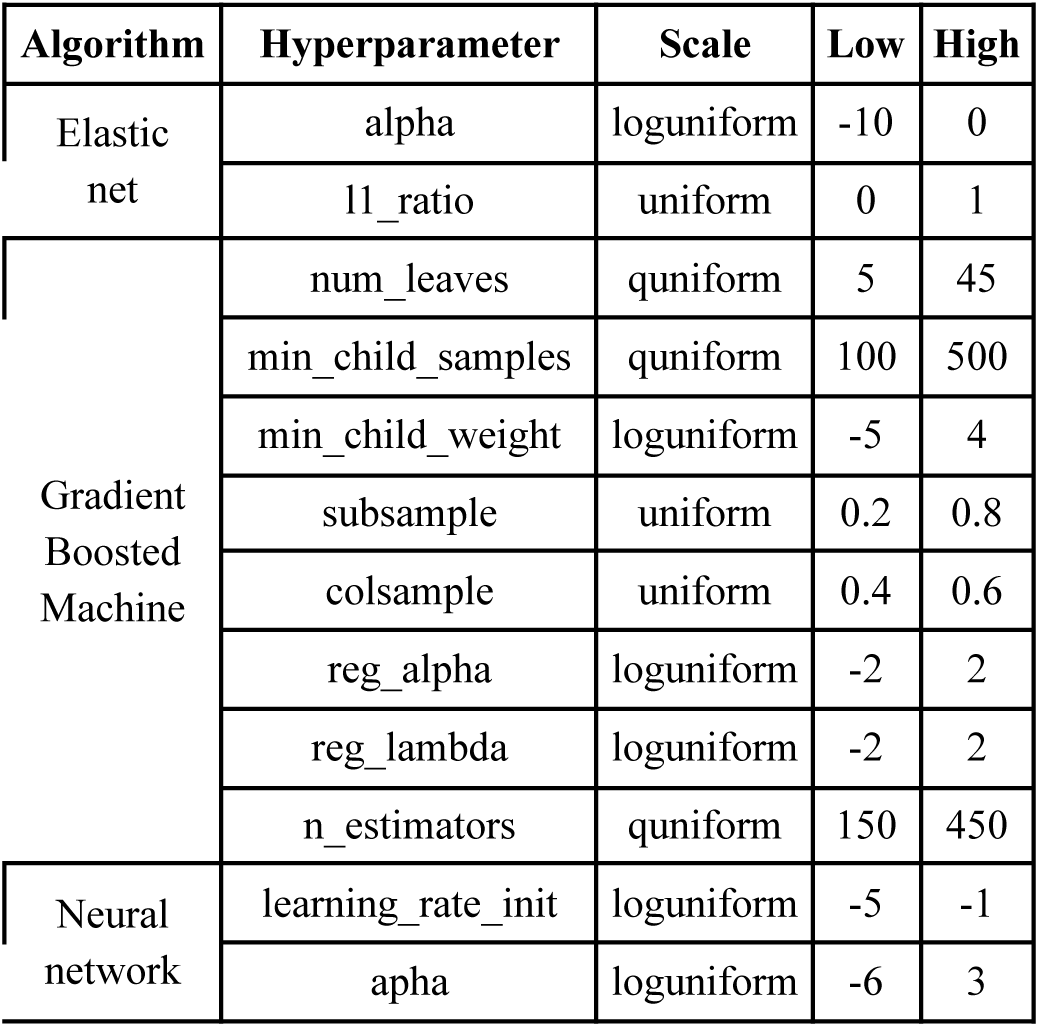
Hyperparameter space for scalar features-based models Bayesian optimization.

**Table S33:**
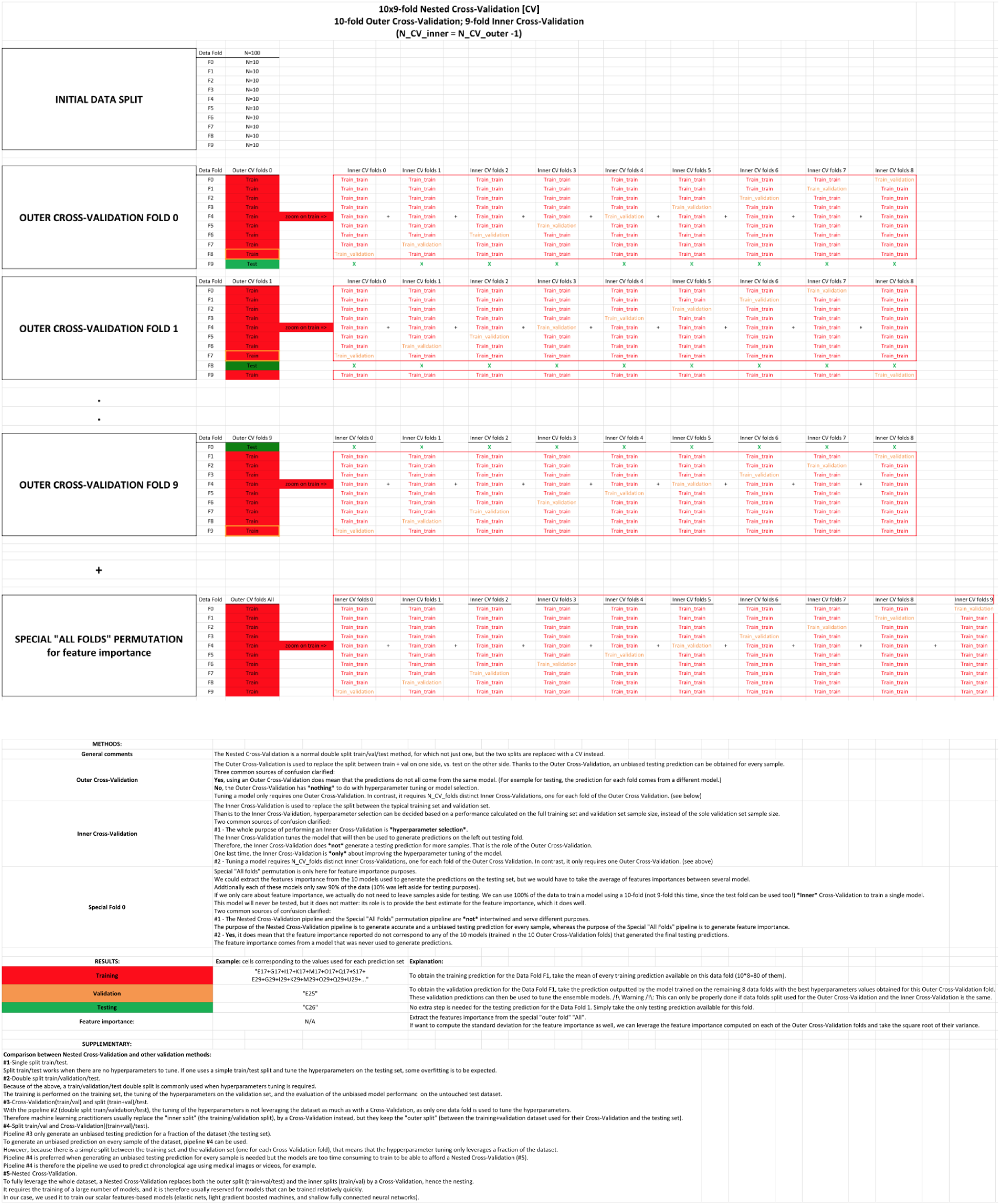
Nested Cross-Validation pipeline.

Table S34: **Hyperparameters tuning upstream of the cross-validation for images-based models**

See supplementary data

**Table S35:** Outer Cross-Validation with inner split pipeline. The values displayed are validation RMSE values. Lower values are associated with better hyperparameter tuning. When two values are displayed (value1/value2), the second value corresponds to the training RMSE. The architecture used was InceptionV3, with an initial learning rate of 0.001. The model was trained on the data folds 2-9, and validated on the data fold #0. The data fold #1 was set aside as the testing set and was not used.

| Data Fold | N=100 | Outer CV folds 0 | Outer CV folds 1 | Outer CV folds 2 | Outer CV folds 3 | Outer CV folds 4 | Outer CV folds 5 | Outer CV folds 6 | Outer CV folds 7 | Outer CV folds 8 | Outer CV folds 9 |
| --- | --- | --- | --- | --- | --- | --- | --- | --- | --- | --- | --- |
| F0 | N=10 | Train | Train | Train | Train | Train | Train | Train | Train | Validation | Test |
| F1 | N=10 | Train | Train | Train | Train | Train | Train | Train | Validation | Test | Train |
| F2 | N=10 | Train | Train | Train | Train | Train | Train | Validation | Test | Train | Train |
| F3 | N=10 | Train | Train | Train | Train | Train | Validation | Test | Train | Train | Train |
| F4 | N=10 | Train | Train | Train | Train | Validation | Test | Train | Train | Train | Train |
| F5 | N=10 | Train | Train | Train | Validation | Test | Train | Train | Train | Train | Train |
| F6 | N=10 | Train | Train | Validation | Test | Train | Train | Train | Train | Train | Train |
| F7 | N=10 | Train | Validation | Test | Train | Train | Train | Train | Train | Train | Train |
| F8 | N=10 | Validation | Test | Train | Train | Train | Train | Train | Train | Train | Train |
| F9 | N=10 | Test | Train | Train | Train | Train | Train | Train | Train | Train | Validation |

## Notes

### Competing Interest Statement

The authors have declared no competing interest.

### Funding Statement

NIEHS R00 ES023504
NIEHS R21 ES25052.
NIAID R01 AI127250
NSF 163870
MassCATS, Massachusetts Life Science Center
Sanofi
The funders had no role in the study design or drafting of the manuscript(s).

### Author Declarations

The Harvard internal review board (IRB) deemed the research as non-human subjects research (IRB: IRB16-2145).

### Summary of Updates

GWAS updated to include imputed data and annotations refreshed using software package FUMA.

## References

Abadi M, Agarwal A, Barham P, Brevdo E, Chen Z, Citro C, Corrado GS, Davis A, Dean J, Devin M, Others. 2015. TensorFlow: Large-scale machine learning on heterogeneous systems.

Agarap AF. 2018. Deep Learning using Rectified Linear Units (ReLU). arXiv [csNE].

Alber M, Lapuschkin S, Seegerer P, Hägele M, Schütt KT, Montavon G, Samek W, Müller K-R, Dähne S, Kindermans P-J. 2019. iNNvestigate neural networks. J Mach Learn Res 20:1–8.

Alqaraawi A, Schuessler M, Weiß P, Costanza E, Berthouze N. 2020. Evaluating saliency map explanations for convolutional neural networks: a user studyProceedings of the 25th International Conference on Intelligent User Interfaces, IUI ’20. New York, NY, USA: Association for Computing Machinery. pp. 275–285.

Asano K, Nomura H, Iwano M, Ando F, Niino N, Shimokata H, Miyake Y. 2005. Relationship between astigmatism and aging in middle-aged and elderly japanese. Japanese Journal of Ophthalmology. doi:10.1007/s10384-004-0152-1

Bergstra J, Bengio Y. 2012. Random search for hyper-parameter optimization. J Mach Learn Res 13:281–305.

Bergstra JS, Bardenet R, Bengio Y, Kégl B. 2011. Algorithms for Hyper-Parameter Optimization In: Shawe-Taylor J, Zemel RS, Bartlett PL, Pereira F, Weinberger KQ, editors. Advances in Neural Information Processing Systems 24. Curran Associates, Inc. pp. 2546–2554.

Bergstra J, Yamins D, Cox D. 2013a. Making a Science of Model Search: Hyperparameter Optimization in Hundreds of Dimensions for Vision Architectures In: Dasgupta S, McAllester D, editors. Proceedings of Machine Learning Research. Atlanta, Georgia, USA: PMLR. pp. 115–123.

Bergstra J, Yamins D, Cox DD. 2013b. Hyperopt: A python library for optimizing the hyperparameters of machine learning algorithmsProceedings of the 12th Python in Science Conference. Citeseer. p. 20.

Bobrov E, Georgievskaya A, Kiselev K, Sevastopolsky A, Zhavoronkov A, Gurov S, Rudakov K, Del Pilar Bonilla Tobar M, Jaspers S, Clemann S. 2018. PhotoAgeClock: deep learning algorithms for development of non-invasive visual biomarkers of aging. Aging 10:3249–3259.

Bos S, Chug E. n.d. Using weight decay to optimize the generalization ability of a perceptron. Proceedings of International Conference on Neural Networks (ICNN’96). doi:10.1109/icnn.1996.548898

Bottou L, Curtis FE, Nocedal J. 2018. Optimization Methods for Large-Scale Machine Learning. SIAM Rev 60:223–311.

Bowne SJ, Daiger SP, Malone KA, Heckenlively JR, Kennan A, Humphries P, Hughbanks-Wheaton D, Birch DG, Liu Q, Pierce EA, Zuo J, Huang Q, Donovan DD, Sullivan LS. 2003. Characterization of RP1L1, a highly polymorphic paralog of the retinitis pigmentosa 1 (RP1) gene. Mol Vis 9:129–137.

Bradski, G. 2000. The OpenCV library. Dr Dobb’s J Software Tools 25:120–125.

Breiman L. 2001. Random Forests. Mach Learn 45:5–32.

Bulik-Sullivan B, Finucane HK, Anttila V, Gusev A, Day FR, Loh P-R, ReproGen Consortium, Psychiatric Genomics Consortium, Genetic Consortium for Anorexia Nervosa of the Wellcome Trust Case Control Consortium 3, Duncan L, Perry JRB, Patterson N, Robinson EB, Daly MJ, Price AL, Neale BM. 2015. An atlas of genetic correlations across human diseases and traits. Nat Genet 47:1236–1241.

Bulik-Sullivan BK, Loh P-R, Finucane HK, Ripke S, Yang J, Patterson N, Daly MJ, Price AL, Neale BM. 2015. LD Score regression distinguishes confounding from polygenicity in genome-wide association studies. Nat Genet 47:291–295.

Bycroft C, Freeman C, Petkova D, Band G, Elliott LT, Sharp K, Motyer A, Vukcevic D, Delaneau O, O’Connell J, Others. 2017. Genome-wide genetic data on\ 500,000 UK Biobank participants. BioRxiv 166298.

Canny J. 1986. A computational approach to edge detection. IEEE Trans Pattern Anal Mach Intell 8:679–698.

Chen T, Guestrin C. 2016. XGBoost: A Scalable Tree Boosting SystemProceedings of the 22nd ACM SIGKDD International Conference on Knowledge Discovery and Data Mining, KDD ’16. New York, NY, USA: Association for Computing Machinery. pp. 785–794.

Chollet F, Others. 2015. keras.

Chueh K-M, Hsieh Y-T, Chen HH, Ma I-H, Huang S-L. n.d. Prediction of Sex and Age from Macular Optical Coherence Tomography Images and Feature Analysis Using Deep Learning. doi:10.1101/2020.12.23.20248805

Clark A. 2018. Pillow Python Imaging Library. Pillow—Pillow (PIL Fork) 5 4 1 documentation.

Consortium T 1000 GP, The 1000 Genomes Project Consortium. 2015. A global reference for human genetic variation. Nature. doi:10.1038/nature15393

Defaye M, Iftinca MC, Gadotti VM, Basso L, Abdullah NS, Cuménal M, Agosti F, Hassan A, Flynn R, Martin J, Soubeyre V, Poulen G, Lonjon N, Vachiery-Lahaye F, Bauchet L, Mery PF, Bourinet E, Zamponi GW, Altier C. 2022. The neuronal tyrosine kinase receptor ligand ALKAL2 mediates persistent pain. J Clin Invest 132. doi:10.1172/JCI154317

De La Cruz N, Shabaneh O, Appiah D. 2021. The Association of Ideal Cardiovascular Health and Ocular Diseases Among US Adults. Am J Med 134:252–259.e1.

de Magalhães JP, Stevens M, Thornton D. 2017. The Business of Anti-Aging Science. Trends Biotechnol 35:1062–1073.

Deng J, Dong W, Socher R, Li L, Kai Li, Li Fei-Fei. 2009. ImageNet: A large-scale hierarchical image database2009 IEEE Conference on Computer Vision and Pattern Recognition. pp. 248–255.

Dhillon S, Shapiro CM, Flanagan J. 2007. Sleep-disordered breathing and effects on ocular health. Can J Ophthalmol 42:238–243.

Duffy BA, Liu M, Flynn T, Toga AW, James Barkovich A, Xu D, Kim H. 2019. Regression activation mapping on the cortical surface using graph convolutional networks.

Duh EJ, Sun JK, Stitt AW. 2017. Diabetic retinopathy: current understanding, mechanisms, and treatment strategies. JCI Insight 2. doi:10.1172/jci.insight.93751

Duke Clinical Research Institute, Elysium Health. 2019. Biomarker Study to Evaluate Correlations Between Epigenetic Aging and NAD+ Levels in Healthy Volunteers.

Ege BM, Hejlesen OK, Larsen OV, Bek T. 2002. The relationship between age and colour content in fundus images. Acta Ophthalmol Scand 80:485–489.

Erbilek M, Fairhurst M, M C D. 2013. Age Prediction from Iris Biometrics. 5th International Conference on Imaging for Crime Detection and Prevention (ICDP 2013). doi:10.1049/ic.2013.0258

EyePACS LLC. 2018. Welcome to EyePACS.

Farrer LA, Adrienne Cupples L, Haines JL, Hyman B, Kukull WA, Mayeux R, Myers RH, Pericak-Vance MA, Risch N, van Duijn CM. 1997. Effects of Age, Sex, and Ethnicity on the Association Between Apolipoprotein E Genotype and Alzheimer Disease: A Meta-analysis. JAMA 278:1349–1356.

Friedman JH. 2001. Greedy Function Approximation: A Gradient Boosting Machine. Ann Stat 29:1189–1232.

Fritsche LG, Igl W, Cooke Bailey JN, Grassmann F, Sengupta S, Bragg-Gresham JL, Burdon KP, Hebbring SJ, Wen C, Gorski M, Kim IK, Cho D, Zack D, Souied E, Scholl HPN, Bala E, Lee KE, Hunter DJ, Sardell RJ, Mitchell P, Merriam JE, Cipriani V, Hoffman JD, Schick T, Lechanteur YTE, Guymer RH, Johnson MP, Jiang Y, Stanton CM, Buitendijk GHS, Zhan X, Kwong AM, Boleda A, Brooks M, Gieser L, Ratnapriya R, Branham KE, Foerster JR, Heckenlively JR, Othman MI, Vote BJ, Liang HH, Souzeau E, McAllister IL, Isaacs T, Hall J, Lake S, Mackey DA, Constable IJ, Craig JE, Kitchner TE, Yang Z, Su Z, Luo H, Chen D, Ouyang H, Flagg K, Lin D, Mao G, Ferreyra H, Stark K, von Strachwitz CN, Wolf A, Brandl C, Rudolph G, Olden M, Morrison MA, Morgan DJ, Schu M, Ahn J, Silvestri G, Tsironi EE, Park KH, Farrer LA, Orlin A, Brucker A, Li M, Curcio CA, Mohand-Saïd S, Sahel J-A, Audo I, Benchaboune M, Cree AJ, Rennie CA, Goverdhan SV, Grunin M, Hagbi-Levi S, Campochiaro P, Katsanis N, Holz FG, Blond F, Blanché H, Deleuze J-F, Igo RP, Truitt B, Peachey NS, Meuer SM, Myers CE, Moore EL, Klein R, Hauser MA, Postel EA, Courtenay MD, Schwartz SG, Kovach JL, Scott WK, Liew G, Tan AG, Gopinath B, Merriam JC, Theodore Smith R, Khan JC, Shahid H, Moore AT, Allie McGrath J, Laux R, Brantley MA, Agarwal A, Ersoy L, Caramoy A, Langmann T, Saksens NTM, de Jong EK, Hoyng CB, Cain MS, Richardson AJ, Martin TM, Blangero J, Weeks DE, Dhillon B, van Duijn CM, Doheny KF, Romm J, Klaver CCW, Hayward C, Gorin MB, Klein ML, Baird PN, den Hollander AI, Fauser S, Yates JRW, Allikmets R, Wang JJ, Schaumberg DA, Klein BEK, Hagstrom SA, Chowers I, Lotery AJ, Léveillard T, Zhang K, Brilliant MH, Hewitt AW, Swaroop A, Chew EY, Pericak-Vance MA, DeAngelis M, Stambolian D, Haines JL, Iyengar SK, Weber BHF, Abecasis GR, Heid IM. 2015. A large genome-wide association study of age-related macular degeneration highlights contributions of rare and common variants. Nat Genet 48:134–143.

Galor A, Lee DJ. 2011. Effects of smoking on ocular health. Curr Opin Ophthalmol 22:477–482.

Glasser A, Croft MA, Kaufman PL. 2001. Aging of the human crystalline lens and presbyopia. Int Ophthalmol Clin 41:1–15.

Gowrisankaran S, Sheedy JE. 2015. Computer vision syndrome: A review. Work 52:303–314.

Grossniklaus HE, Nickerson JM, Edelhauser HF, Bergman LAMK, Berglin L. 2013. Anatomic alterations in aging and age-related diseases of the eye. Invest Ophthalmol Vis Sci 54:ORSF23–7.

Guan J, Umapathy G, Yamazaki Y, Wolfstetter G, Mendoza P, Pfeifer K, Mohammed A, Hugosson F, Zhang H, Hsu AW, Halenbeck R, Hallberg B, Palmer RH. 2015. FAM150A and FAM150B are activating ligands for anaplastic lymphoma kinase. Elife 4:e09811.

Hayashi K, Masumoto M, Fujino S, Hayashi F. 1993. [Changes in corneal astigmatism with aging]. Nihon Ganka Gakkai Zasshi 97:1193–1196.

Hinton G. n.d. Slide 29 of Lecture 6, Geoffrey Hinton coursera’s class. http://www.cs.toronto.edu. http://www.cs.toronto.edu/~tijmen/csc321/slides/lecture_slides_lec6.pdf

Hochreiter S. 1991. Untersuchungen zu dynamischen neuronalen Netzen. Diploma, Technische Universität München 91.

Hochreiter S, Bengio Y, Frasconi P, Schmidhuber J, Others. 2001. Gradient flow in recurrent nets: the difficulty of learning long-term dependencies.

Hoerl AE, Kennard RW. 1970. Ridge Regression: Biased Estimation for Nonorthogonal Problems. null 12:55–67.

Ho J-D, Liou S-W, Tsai RJ-F, Tsai C-Y. 2010. Effects of aging on anterior and posterior corneal astigmatism. Cornea 29:632–637.

Horvath S. 2013. DNA methylation age of human tissues and cell types. Genome Biol 14:R115.

Horvath S, Erhart W, Brosch M, Ammerpohl O, von Schönfels W, Ahrens M, Heits N, Bell JT, Tsai P-C, Spector TD, Deloukas P, Siebert R, Sipos B, Becker T, Röcken C, Schafmayer C, Hampe J. 2014. Obesity accelerates epigenetic aging of human liver. Proc Natl Acad Sci U S A 111:15538–15543.

Hossain S, Calloway C, Lippa D, Niederhut D, Shupe D. 2019. Visualization of Bioinformatics Data with Dash BioProceedings of the 18th Python in Science Conference. pp. 126–133.

Hunter JD. 2007. Matplotlib: A 2D Graphics Environment. Comput Sci Eng 9:90–95.

Inc PT. 2015. Collaborative data science. Montreal: Plotly Technologies Inc Montral.

Ke G, Meng Q, Finley T, Wang T, Chen W, Ma W, Ye Q, Liu T-Y. 2017. LightGBM: A Highly Efficient Gradient Boosting Decision Tree In: Guyon I, Luxburg UV, Bengio S, Wallach H, Fergus R, Vishwanathan S, Garnett R, editors. Advances in Neural Information Processing Systems 30. Curran Associates, Inc. pp. 3146–3154.

Kim C-S, Kim M-Y, Kim H-S, Lee Y-C. 2002. Change of Corneal Astigmatism with Aging in Koreans with Normal Visual Acuity. Journal of The Korean Ophthalmological Society 43:1956–1962.

Kim J-M. 2000. The Effects of Drugs, including Alcohol, on Ocular Health and Contact Lens Wear. Journal of Korean Ophthalmic Optics Society 5:73–81.

Kingma DP, Ba J. 2014. Adam: A Method for Stochastic Optimization. arXiv [csLG].

Klambauer G, Unterthiner T, Mayr A, Hochreiter S. 2017. Self-Normalizing Neural Networks In: Guyon I, Luxburg UV, Bengio S, Wallach H, Fergus R, Vishwanathan S, Garnett R, editors. Advances in Neural Information Processing Systems 30. Curran Associates, Inc. pp. 971–980.

Kohavi R, Others. 1995. A study of cross-validation and bootstrap for accuracy estimation and model selectionIjcai. Montreal, Canada. pp. 1137–1145.

Kotikalapudi R, Others. 2019. keras-vis. 2017. URL https://github com/raghakot/keras-vis.

Krizhevsky A, Sutskever I, Hinton GE. 2012. ImageNet Classification with Deep Convolutional Neural Networks In: Pereira F, Burges CJC, Bottou L, Weinberger KQ, editors. Advances in Neural Information Processing Systems 25. Curran Associates, Inc. pp. 1097–1105.

Krogh A, Hertz JA. 1992. A Simple Weight Decay Can Improve Generalization In: Moody JE, Hanson SJ, Lippmann RP, editors. Advances in Neural Information Processing Systems 4. Morgan-Kaufmann. pp. 950–957.

LeCun Y, Bengio Y, Hinton G. 2015. Deep learning. Nature. doi:10.1038/nature14539

Le Goallec A, Collin S, Diai S, Prost J-B, Jabri M ’hamed, Vincent T, Patel CJ. 2021. Analyzing the multidimensionality of biological aging with the tools of deep learning across diverse image-based and physiological indicators yields robust age predictors. medRxiv.

Le Goallec A, Diai S, Collin S, Prost J-B, Vincent T, Patel CJ. 2022. Using deep learning to predict abdominal age from liver and pancreas magnetic resonance images. Nat Commun 13:1979.

Le Goallec A, Patel CJ. 2019. Age-dependent co-dependency structure of biomarkers in the general population of the United States. Aging 11:1404–1426.

Li X, Ploner A, Wang Y, Magnusson PKE, Reynolds C, Finkel D, Pedersen NL, Jylhävä J, Hägg S. 2020. Longitudinal trajectories, correlations and mortality associations of nine biological ages across 20-years follow-up. eLife. doi:10.7554/elife.51507

Loh P-R, Kichaev G, Gazal S, Schoech AP, Price AL. 2018. Mixed-model association for biobank-scale datasets. Nat Genet 50:906–908.

Loh P-R, Schizophrenia Working Group of the Psychiatric Genomics Consortium, Bhatia G, Gusev A, Finucane HK, Bulik-Sullivan BK, Pollack SJ, de Candia TR, Lee SH, Wray NR, Kendler KS, O’Donovan MC, Neale BM, Patterson N, Price AL. 2015a. Contrasting genetic architectures of schizophrenia and other complex diseases using fast variance-components analysis. Nature Genetics. doi:10.1038/ng.3431

Loh P-R, Tucker G, Bulik-Sullivan BK, Vilhjálmsson BJ, Finucane HK, Salem RM, Chasman DI, Ridker PM, Neale BM, Berger B, Patterson N, Price AL. 2015b. Efficient Bayesian mixed-model analysis increases association power in large cohorts. Nat Genet 47:284–290.

Lord SR, Smith ST, Menant JC. 2010. Vision and falls in older people: risk factors and intervention strategies. Clin Geriatr Med 26:569–581.

McKinney W, Others. 2010. Data structures for statistical computing in pythonProceedings of the 9th Python in Science Conference. Austin, TX. pp. 51–56.

Millman KJ, Jarrod Millman K, Aivazis M. 2011. Python for Scientists and Engineers. Computing in Science & Engineering. doi:10.1109/mcse.2011.36

Nagasato D, Tabuchi H, Masumoto H, Kusuyama T, Kawai Y, Ishitobi N, Furukawa H, Adachi S, Murao F, Mitamura Y. 2020. Prediction of age and brachial-ankle pulse-wave velocity using ultra-wide-field pseudo-color images by deep learning. Sci Rep 10:19369.

Nair V, Hinton GE. 2010. Rectified Linear Units Improve Restricted Boltzmann Machines.

Ohia SE, Njie-Mbye YF, Opere CA, Kulkarni M, Barett A. 2014. Chapter 22 - Ocular Health, Vision, and a Healthy Diet In: Rahman I, Bagchi D, editors. Inflammation, Advancing Age and Nutrition. San Diego: Academic Press. pp. 267–277.

Oliphant TE. 2007. Python for Scientific Computing. Computing in Science Engineering 9:10–20.

Oliphant TE. 2006. A guide to NumPy. Trelgol Publishing USA.

Ong SR, Crowston JG, Loprinzi PD, Ramulu PY. 2018. Physical activity, visual impairment, and eye disease. Eye 32:1296–1303.

Pan SJ, Yang Q. 2010. A Survey on Transfer Learning. IEEE Trans Knowl Data Eng 22:1345–1359.

Patel CJ, Bhattacharya J, Butte AJ. 2010. An Environment-Wide Association Study (EWAS) on type 2 diabetes mellitus. PLoS One 5:e10746.

Pedregosa F, Varoquaux G, Gramfort A, Michel V, Thirion B, Grisel O, Blondel M, Prettenhofer P, Weiss R, Dubourg V, Others. 2011. Scikit-learn: Machine learning in Python. the Journal of machine Learning research 12:2825–2830.

Popescu M-C, Balas VE, Perescu-Popescu L, Mastorakis N. 2009. Multilayer perceptron and neural networks. WSEAS Trans Circuits and Syst 8.

Poplin R, Varadarajan AV, Blumer K, Liu Y, McConnell MV, Corrado GS, Peng L, Webster DR. 2018. Prediction of cardiovascular risk factors from retinal fundus photographs via deep learning. Nat Biomed Eng 2:158–164.

Poplin R, Varadarajan AV, Blumer K, Liu Y, Mc Connell MV, Corrado GS, Peng L, Webster DR. n.d. Predicting Cardiovascular Risk Factors from Retinal Fundus Photographs using Deep Learning.

Prechelt L. 1998. Early Stopping - But When? In: Orr GB, Müller K-R, editors. Neural Networks: Tricks of the Trade. Berlin, Heidelberg: Springer Berlin Heidelberg. pp. 55–69.

Rajput M, Sable G. n.d. Deep Learning Based Gender and Age Estimation from Human Iris. SSRN Electronic Journal. doi:10.2139/ssrn.3576471

Rein DB, Zhang P, Wirth KE, Lee PP, Hoerger TJ, McCall N, Klein R, Tielsch JM, Vijan S, Saaddine J. 2006. The economic burden of major adult visual disorders in the United States. Arch Ophthalmol 124:1754–1760.

Reshetnyak AV, Murray PB, Shi X, Mo ES, Mohanty J, Tome F, Bai H, Gunel M, Lax I, Schlessinger J. 2015. Augmentor α and β (FAM150) are ligands of the receptor tyrosine kinases ALK and LTK: Hierarchy and specificity of ligand-receptor interactions. Proc Natl Acad Sci U S A 112:15862–15867.

Ribeiro MT, Singh S, Guestrin C. 2016. “ Why should I trust you?” Explaining the predictions of any classifierProceedings of the 22nd ACM SIGKDD International Conference on Knowledge Discovery and Data Mining. pp. 1135–1144.

Rosenblatt F. 1958. The Perceptron: A Theory of Statistical Separability in Cognitive Systems (Project Para). Cornell Aeronautical Laboratory.

Ruder S. 2016. An overview of gradient descent optimization algorithms. arXiv [csLG].

Russakovsky O, Deng J, Su H, Krause J, Satheesh S, Ma S, Huang Z, Karpathy A, Khosla A, Bernstein M, Berg AC, Fei-Fei L. 2015. ImageNet Large Scale Visual Recognition Challenge. Int J Comput Vis 115:211–252.

Salvi SM, Akhtar S, Currie Z. 2006. Ageing changes in the eye. Postgrad Med J 82:581–587.

Selvaraju RR, Cogswell M, Das A, Vedantam R, Parikh D, Batra D. 2017. Grad-cam: Visual explanations from deep networks via gradient-based localizationProceedings of the IEEE International Conference on Computer Vision. pp. 618–626.

Sgroi A, Bowyer KW, Flynn PJ. 2013. The prediction of old and young subjects from iris texture2013 International Conference on Biometrics (ICB). pp. 1–5.

Shigueoka LS, Mariottoni EB, Thompson AC, Jammal AA, Costa VP, Medeiros FA. 2021. Predicting Age From Optical Coherence Tomography Scans With Deep Learning. Transl Vis Sci Technol 10:12.

Shorten C, Khoshgoftaar TM. 2019. A survey on Image Data Augmentation for Deep Learning. Journal of Big Data 6:60.

Simonyan K, Zisserman A. 2014. Very Deep Convolutional Networks for Large-Scale Image Recognition. arXiv [csCV].

Sim YS, Yang SW, Park YL, Na KS, Kim HS. 2017. Age-related Changes in Anterior, Posterior Corneal Astigmatism in a Korean Population. Journal of the Korean Ophthalmological Society. doi:10.3341/jkos.2017.58.8.911

Sparkes RS, Heinzmann C, Goldflam S, Kojis T, Saari JC, Mohandas T, Klisak I, Bateman JB, Crabb JW. 1992. Assignment of the gene (RLBP1) for cellular retinaldehyde-binding protein (CRALBP) to human chromosome 15q26 and mouse chromosome 7. Genomics 12:58–62.

Srivastava N, Hinton G, Krizhevsky A, Sutskever I, Salakhutdinov R. 2014. Dropout: a simple way to prevent neural networks from overfitting. J Mach Learn Res 15:1929–1958.

Sudlow C, Gallacher J, Allen N, Beral V, Burton P, Danesh J, Downey P, Elliott P, Green J, Landray M, Liu B, Matthews P, Ong G, Pell J, Silman A, Young A, Sprosen T, Peakman T, Collins R. 2015. UK biobank: an open access resource for identifying the causes of a wide range of complex diseases of middle and old age. PLoS Med 12:e1001779.

Szegedy C, Ioffe S, Vanhoucke V, Alemi AA. 2017. Inception-v4, inception-resnet and the impact of residual connections on learningThirty-First AAAI Conference on Artificial Intelligence.

Szegedy C, Vanhoucke V, Ioffe S, Shlens J, Wojna Z. 2016. Rethinking the inception architecture for computer visionProceedings of the IEEE Conference on Computer Vision and Pattern Recognition. pp. 2818–2826.

Tan C, Sun F, Kong T, Zhang W, Yang C, Liu C. 2018. A Survey on Deep Transfer LearningArtificial Neural Networks and Machine Learning – ICANN 2018. Springer International Publishing. pp. 270–279.

Tan M, Le QV. 2019. EfficientNet: Rethinking Model Scaling for Convolutional Neural Networks. arXiv [csLG].

Tibshirani R. 1996. Regression shrinkage and selection via the lasso. J R Stat Soc Series B Stat Methodol 58:267–288.

Ueno Y, Hiraoka T, Beheregaray S, Miyazaki M, Ito M, Oshika T. 2014. Age-related changes in anterior, posterior, and total corneal astigmatism. J Refract Surg 30:192–197.

Van Rossum G, Drake FL. 2011. The Python Language Reference Manual. Network Theory Limited.

Vavvas DG, Small KW, Awh CC, Zanke BW, Tibshirani RJ, Kustra R. 2018. *CFH* and *ARMS2* genetic risk determines progression to neovascular age-related macular degeneration after antioxidant and zinc supplementation. Proc Natl Acad Sci U S A 115:E696–E704.

Virtanen P, SciPy 1. 0 Contributors, Gommers R, Oliphant TE, Haberland M, Reddy T, Cournapeau D, Burovski E, Peterson P, Weckesser W, Bright J, van der Walt SJ, Brett M, Wilson J, Jarrod Millman K, Mayorov N, Nelson ARJ, Jones E, Kern R, Larson E, Carey CJ, Polat İ, Feng Y, Moore EW, VanderPlas J, Laxalde D, Perktold J, Cimrman R, Henriksen I, Quintero EA, Harris CR, Archibald AM, Ribeiro AH, Pedregosa F, van Mulbregt P. 2020. SciPy 1.0: fundamental algorithms for scientific computing in Python. Nature Methods. doi:10.1038/s41592-019-0686-2

Visser L. 2006. Common eye disorders in the elderly—a short review. South African Family Practice. doi:10.1080/20786204.2006.10873425

Walt S van der, van der Walt S, Chris Colbert S, Varoquaux G. 2011. The NumPy Array: A Structure for Efficient Numerical Computation. Computing in Science & Engineering. doi:10.1109/mcse.2011.37

Wang K, Li M, Hakonarson H. 2010. ANNOVAR: functional annotation of genetic variants from high-throughput sequencing data. Nucleic Acids Res 38:e164.

Wang Z, Yang J. 2017. Diabetic Retinopathy Detection via Deep Convolutional Networks for Discriminative Localization and Visual Explanation. arXiv [csCV].

Watanabe K, Taskesen E, van Bochoven A, Posthuma D. 2017. Functional mapping and annotation of genetic associations with FUMA. Nat Commun 8:1826.

Weiss K, Khoshgoftaar TM, Wang D. 2016. A survey of transfer learning. Journal of Big data 3:9.

Winkler TW, Grassmann F, Brandl C, Kiel C, Günther F, Strunz T, Weidner L, Zimmermann ME, Korb CA, Poplawski A, Schuster AK, Müller-Nurasyid M, Peters A, Rauscher FG, Elze T, Horn K, Scholz M, Cañadas-Garre M, McKnight AJ, Quinn N, Hogg RE, Küchenhoff H, Heid IM, Stark KJ, Weber BHF. 2020. Genome-wide association meta-analysis for early age-related macular degeneration highlights novel loci and insights for advanced disease. BMC Med Genomics 13:120.

Zeiler MD. 2012. ADADELTA: An Adaptive Learning Rate Method. arXiv [csLG].

Zhang J, He T, Sra S, Jadbabaie A. 2019. Why gradient clipping accelerates training: A theoretical justification for adaptivity. arXiv [mathOC].

Zhou B, Khosla A, Lapedriza A, Oliva A, Torralba A. 2016. Learning deep features for discriminative localizationProceedings of the IEEE Conference on Computer Vision and Pattern Recognition. pp. 2921–2929.

Zou H, Hastie T. 2005. Regularization and variable selection via the elastic net. J R Stat Soc Series B Stat Methodol 67:301–320.

